# Multi-cohort transcriptomic subtyping of B-cell acute lymphoblastic leukemia

**DOI:** 10.1101/2022.02.17.22270919

**Authors:** Ville-Petteri Mäkinen, Jacqueline Rehn, James Breen, David Yeung, Deborah L White

## Abstract

Acute lymphoblastic leukemia (ALL) is the most common childhood cancer and comprises multiple genetically distinguishable subtypes. To detect subtypes, current pipelines include fusion calling, polymorphisms, candidate gene copy numbers and cytogenetics but these approaches have limitations. RNA-seq provides a functional genome-wide snapshot that enables classification of ALL subtypes, however, typical mRNA-seq clustering analyses lack the rigor of quantitative modelling. Furthermore, high-dimensional gene expression data across cohorts and countries contain biases that previous transcriptomics studies have not addressed. Our aim was to integrate easy-to-interpret reliable transcriptome-wide biomarkers into subtyping pipelines. We analyzed 2,046 samples from two continents, carefully adjusted for biases and applied a rigorous machine learning design with independent replication. Six ALL subtypes that covered 32% of patients were robustly detected by mRNA-seq (PPV ≥ 87%). Five other frequent subtypes were distinguishable in 40% of patients, although overlapping transcriptional profiles led to lower accuracy (52% ≤ PPV ≤ 73%). Based on these findings, we developed the Allspice tool that predicts ALL subtypes and driver genes from unadjusted mRNA-seq read counts as encountered in real-world settings. Allspice also includes quantitative classification and safety metrics to help determine the most plausible genetic drivers for cases where other findings are inconclusive.

## Background

Acute lymphoblastic leukemia (ALL) is characterized by abnormal differentiation and proliferation of malignant lymphoid precursors in blood and bone marrow [1, 2]. Usually, the disease manifests as abnormal proliferation of B-cells while less than a quarter of patients present with a T-cell malignancy. The incidence rate is the highest in children under the age of 10 and in adults over the age of 65 with an average age-adjusted global annual incidence of 0.85 per 100,000 individuals [3]. Despite recent progress in molecular phenotyping, ALL remains a life-threatening disease, and advanced age (beyond paediatric) and certain subtypes are predictive of poor outcomes [1,2,4–7]. For these reasons, there is a compelling rationale for improving diagnostic tools and for pursuing deeper biological insight into ALL through new emerging technologies.

Cytogenetic testing, immunophenotyping and molecular assays are essential for the diagnosis and further stratification of the disease into subtypes with different biological characteristics and prognosis [8–10]. Transcriptomic profiling of lymphoblastic cells is a recent addition to the diagnostic toolbox and it has led to the detection of new ALL subtypes [11–15]. Standard cytogenetic testing (G-banded karyotype) with adjunctive fluorescence in situ hybridization (FISH) can detect aneuploidy and chromosomal translocations such as *BCR-ABL1*, *ETV6-RUNX1* and *TCF3-PBX1* fusions [8, 16]. *KMT2A* lesions, intrachromosomal amplification of chromosome 21 (iAMP21) and *IGH-CRLF2* and *P2RY8-CRLF2* fusions are also detectable. Other subtypes are identified only through additional analysis such as real-time PCR, single nucleotide polymorphisms and RNA sequencing. Subtypes such as *DUX4* rearrangements, *ETV6-RUNX1*-like and *PAX5* alterations are examples where the underlying genomic alterations are difficult to detect using standard of care laboratory methods and RNA-seq profiling has emerged as an important diagnostic support [11,13,17]. For these reasons, understanding the information carried by transcriptomes in relation to the genome alterations in ALL is important, particularly for those patients for whom a conclusive genomic driver cannot be determined by current molecular diagnostics.

The high volume of RNA sequencing data per individual makes it necessary to employ machine learning techniques to process the raw information for the identification of subtypes [7,11,12,18– 20]. In one such study, Gu et al. used clustering algorithms to characterize the transcriptional landscape of ALL [12]; the clusters were further investigated against identifiable genomic lesions to classify individual patients and to refine the taxonomy of ALL. Using the taxonomy, the authors were able to assign subtypes for 94% of the study subjects. Recently, Schmidt et al. used the taxonomy to classify patients in multiple datasets [11, 21]. Encouraged by these successes, we set out to leverage gene expression data within our transcriptomic analysis pipeline of B-cell ALL to improve diagnoses and to gain biological insight into the transcriptional landscape of ALL.

Several issues related to mRNA-seq profiling of ALL have not yet been addressed. Firstly, clustering analyses of mRNA-seq data should be subjected to the rigorous adjustment of biases that is standard practice in epidemiology [22–24], instead, most studies opt to (mis)use methods such as surrogate variable analysis that may cause artefacts if the data batches and biological characteristics are correlated (as they tend to be in most multi-cohort collections). These artefacts will be further amplified by clustering algorithms and machine learning models. Cohort biases may explain why gene signatures of subtypes derived from gene expression clusters such as Ph-like have been difficult to consolidate between different studies without additional experiments and analyses [25–27].

Secondly, a taxonomy that is based on gene expression profiles should not be used when fitting a machine learning model to RNA-seq read counts. Under the worst-case scenario, a transcriptome dataset affected by cohort bias leads to artificial clustering; the same artificial clustering of gene expression is then captured by machine learning and passed on as a falsely distinct subtype. Instead, we propose that gene expression classifiers should be trained with directly observable sequence variants or otherwise independently distinguishable subtypes or with longitudinal data on clinical outcomes and treatment effects.

The aim of this study is to introduce a reliable ALL classifier that can be integrated into current transcriptomic analyses with minimal additional resources and that can reliably classify ALL cases in an unbiased manner. To achieve reliability, we use new techniques to adjust for cohort and RNA-seq platform biases in a set of n = 1,279 North American patients and then validate the predictions in an independent cohort of n = 767 Australian patients. To achieve easy integration into existing workflows, we introduce the Allspice R package with extensive documentation, small programmatic footprint and additional features for predicting genomic drivers and for confirming the tissue-identity of the biological samples. The tool is also easy to train for any other disease or classification problem or to update with improved models of ALL in the future.

## Results

### Cohort characteristics and study design

The distribution of ALL subtypes across cohorts is shown in Table 1. Note the definition of genomic subtypes: we excluded categories such as Ph-like due to the technical reasons described in Methods and in Supplement Figure S1. The most common subtypes included *CRLF2* (between 3.0% and 18.4% of cohort participants), *ETV6-RUNX1* (≥9.9% of pediatric patients), Hyperdiploid (≥9.8% of pediatric patients), *KMT2A* (between 2.2% and 13.4%) and Ph+ (between 2.4% and 21.5%). There were differences between the cohorts regarding age (P ≤ 7.6 × 10^−11^) and several subtypes including pediatric *CRLF2* (P = 8.4 × 10^−12^), pediatric *ETV6-RUNX1* (P = 2.3 × 10^−8^), *KMT2A* across both age categories (P ≤ 3.2 × 10^−7^) and non-pediatric Ph+ (P = 0.00016). The percentage of undefined samples was between 8.7% in the St Jude cohort and 33.1% in the Australian paediatric cohort.

**Table 1.** Patient characteristics and frequencies (%) of ALL subtypes according to known genetic alterations. The participants were grouped primarily by the recruiting institute and secondarily by age (>99% of participants in the pediatric cohorts were below 20 years of age). Mean and standard deviation are shown for age. P-values for cohort differences were calculated by the χ^2^-test.

|  | Pediatric cohorts |  |  |  | Adult and general cohorts |  |  |
| --- | --- | --- | --- | --- | --- | --- | --- |
|  | COG | St Jude | Australia | P-value | ECOG-ACRIN,<br>Toronto, MDACC,<br>CALGB | Australia | P-value |
| Male | 177 | 221 | 83 | $4.4 \times 10^{-6}$ | 251 | 304 | 0.021 |
| Female | 120 | 188 | 73 | 0.016 | 233 | 205 | 0.013 |
| Unknown | 27 | 52 | 89 | $1.8 \times 10^{-21}$ | 10 | 13 | 0.77 |
| Age (years) | 9.2 ± 5.7 | 6.4 ± 4.4 | 7.4 ± 4.0 | $7.6 \times 10^{-11}$ | 45.1 ± 14.8 | 35.7 ± 22.6 | $3.6 \times 10^{-13}$ |
| BCL2/MYC | 0.0 | 0.4 | 0.4 | 0.50 | 2.8 | 0.8 | 0.024 |
| CDX2 hi-exp | 0.3 | 0.0 | 0.0 | 0.34 | 0.6 | 0.6 | 1.0 |
| CRLF2 | 7.4 | 3.0 | 18.4 | $8.4 \times 10^{-12}$ | 13.2 | 8.4 | 0.020 |
| DUX4 | 7.4 | 7.4 | 2.0 | 0.0096 | 4.9 | 6.9 | 0.21 |
| ETV6-RUNX1 | 9.9 | 26.7 | 18.0 | $2.3 \times 10^{-8}$ | 1.0 | 2.3 | 0.18 |
| HLF | 0.0 | 0.4 | 0.4 | 0.50 | 0.6 | 0.6 | 1.0 |
| Hyperdiploid | 21.0 | 21.3 | 9.8 | 0.00034 | 4.0 | 5.7 | 0.27 |
| Hypodiploid | 1.2 | 0.7 | 0.8 | 0.68 | 11.5 | 3.6 | $3.1 \times 10^{-6}$ |
| iAMP21 | 6.5 | 0.4 | 0.8 | $0.72 \times 10^{-8}$ | 0.2 | 0.4 | 1.0 |
| IKZF1 N159Y | 0.3 | 0.0 | 1.3 | 0.043 | 0.4 | 0.6 | 1.0 |
| KMT2A | 2.2 | 10.8 | 2.9 | $2.0 \times 10^{-7}$ | 13.4 | 4.4 | $3.2 \times 10^{-7}$ |
| MEF2D | 2.2 | 1.1 | 0.0 | 0.058 | 2.0 | 1.1 | 0.39 |
| NUTM1 | 0.3 | 1.1 | 0.4 | 0.36 | 0.0 | 0.2 | 1.0 |
| PAX5 Alt | 4.0 | 5.9 | 3.7 | 0.32 | 8.3 | 1.9 | $6.4 \times 10^{-6}$ |
| PAX5 P80R | 0.9 | 1.1 | 0.8 | 0.94 | 3.4 | 2.9 | 0.74 |
| Ph+ | 5.6 | 2.4 | 4.1 | 0.070 | 12.3 | 21.5 | $1.6 \times 10^{-4}$ |
| TCF3-PBX1 | 2.2 | 6.7 | 1.2 | 0.00023 | 3.0 | 2.3 | 0.59 |
| ZNF384 | 1.9 | 2.0 | 2.0 | 0.99 | 2.8 | 4.4 | 0.24 |
| Undefined | 27.2 | 8.7 | 33.1 | $1.7 \times 10^{-16}$ | 15.6 | 31.2 | $7.0 \times 10^{-9}$ |

The RNA data were filtered and normalized separately for the North American and Australian samples (Figure 1A,B) and the training data were adjusted for technical and cohort confounders (Figure 1C). We also saved the unadjusted RNA data and compared it against the adjusted data to remove the most confounded genes (Figure 1D). In the next step, we created an internal training and testing set by splitting the North American data into two random subsets (Figure 1E). These two subsets were used as the initial material for selecting features and controlling for the complexity of machine learning models via hyperparameters. For the full models, we used all North American samples for training and the Australian dataset as an independent external validation set (Figure 1F).

**Figure 1.**
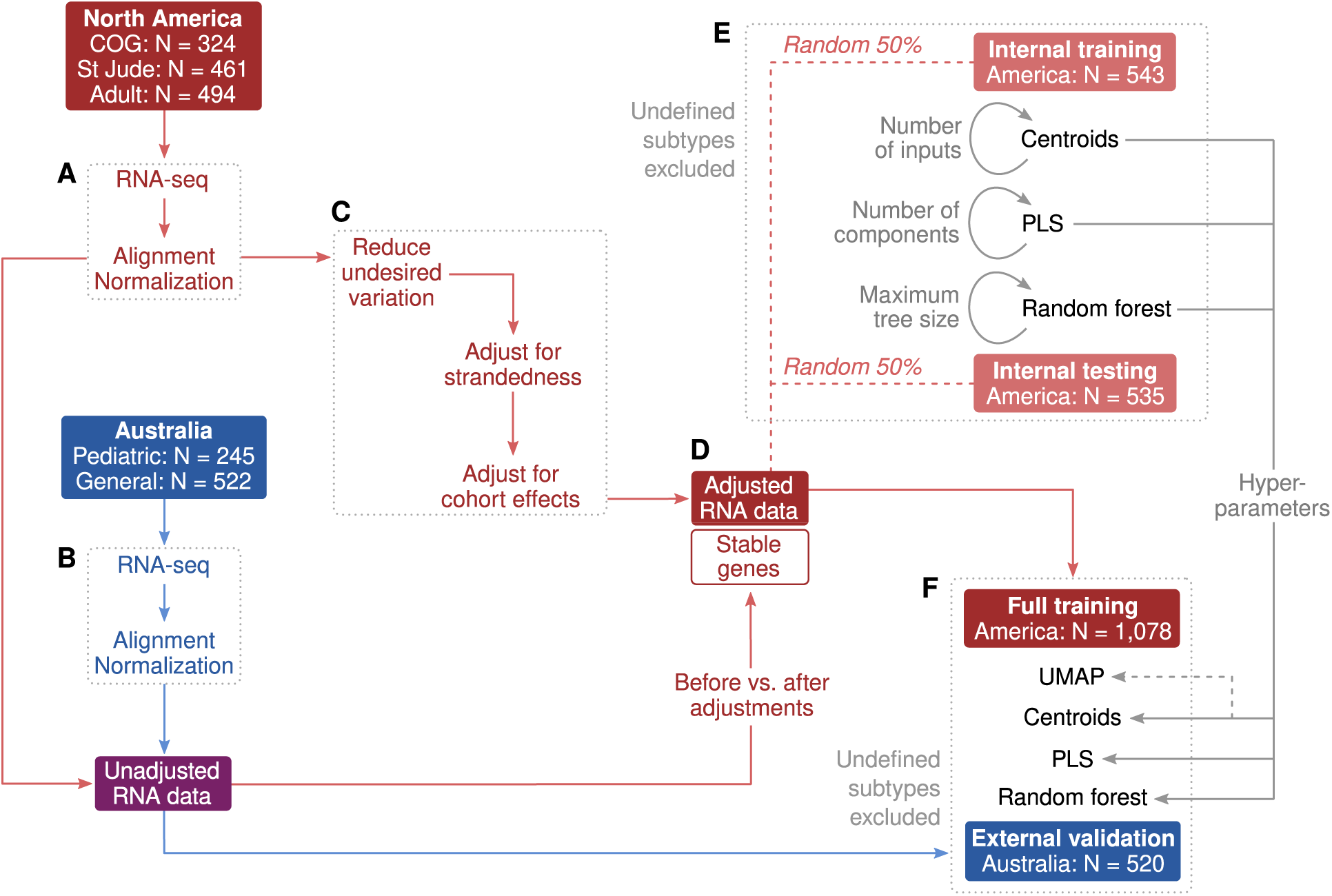
Study design. **A,B**) RNA-seq pre-processing was applied separately for North American and Australian datasets. **C**) Non-biological differences due to technical artifacts and cohort effects were adjusted according to genetically matched subsets (details in Methods). **D**) Correlation coefficients were calculated between unadjusted and adjusted expression levels to exclude genes that were heavily influenced by confounders. A total of 6,673 stable genes with R ≥ 0.9 were included for subtype modelling. **E**) Internal training and testing sets were randomly chosen from the North American participants as initial material to evaluate machine learning models and to determine hyperparameters. The randomization was done separately for each subtype to ensure matching subtype frequencies, which explains the small difference in set sizes. **F**) Final machine learning models were trained with the full North American dataset and validated in the Australian dataset.

### Adjustments for confounders

Confounding factors were mitigated first by surrogate variable analysis within pre-defined cohort batches (details in Methods and in Supplement Figure S2), and batch differences between the cohorts were then adjusted with a new algorithm we recently developed for time-series metabolomics data (manuscript in revision). The adjustments removed correlations between gene expression and RNA library format (Figure 2A,D) and between the two continents of origin (Figure 2B,E) but did not influence the correlations between gene expression and ALL subtypes (Figure 2C,F; Supplement Figures S5-S7).

**Figure 2.**
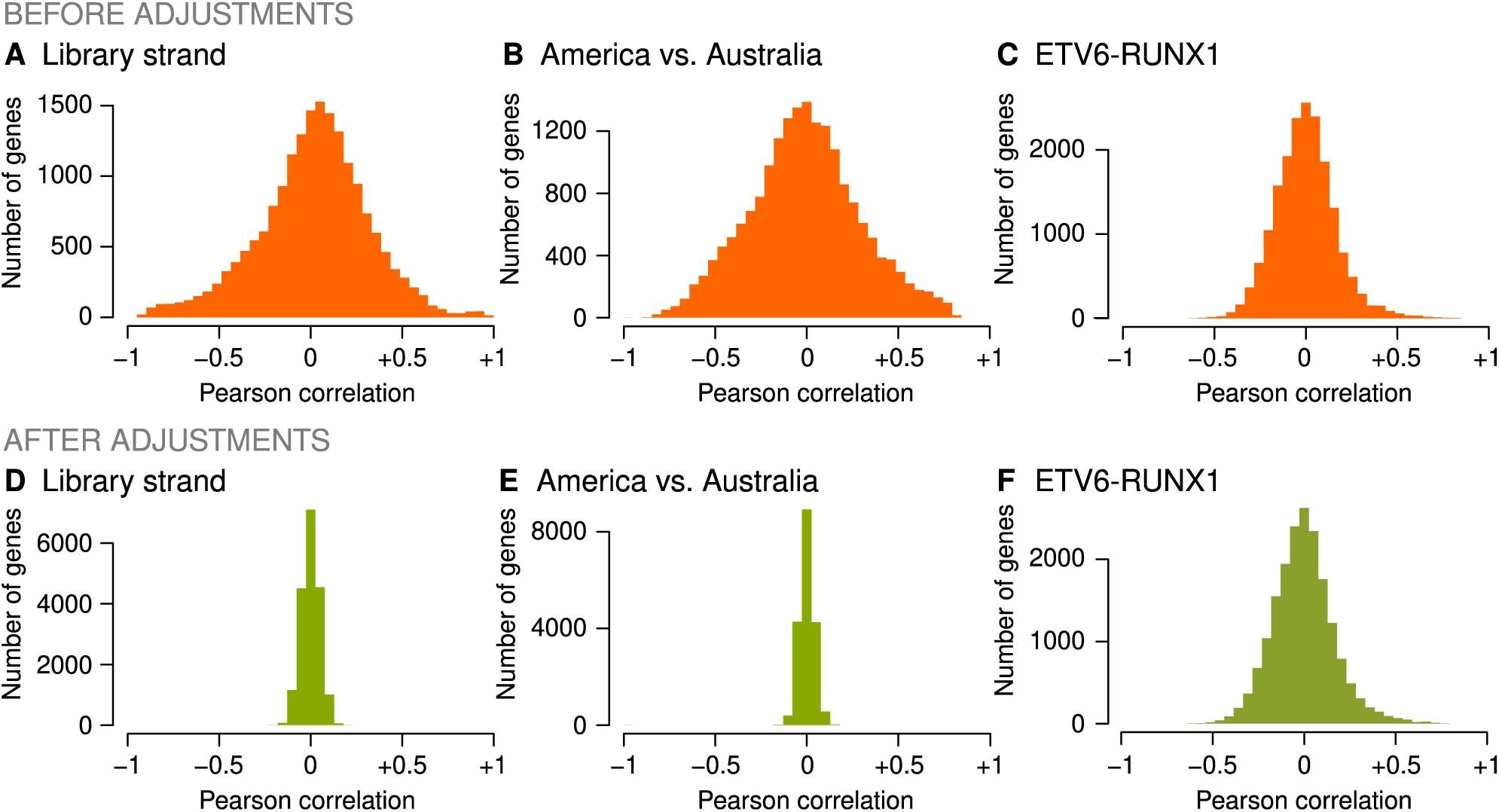
Impact of confounder adjustments. Pearson correlation coefficients were calculated between 18,503 log-transformed genes and a technical or clinical variable before and after gene expression levels were adjusted as described in Methods. A wide histogram indicates substantial covariation across the transcriptome while a narrow histogram indicates successful removal of covariance. **A,D**) The North American cohorts included 204 (16%) RNA samples that were sequenced with an unstranded library. **B,E**) Strandedness and other technical differences manifested as substantial co-variation between the transcriptome and the continent of origin. **C,F**) Covariation between gene expression and ALL subtypes such as ETV6-RUNX1 was preserved.

To verify that the data processing was technically sound, we constructed a visual layout of the individuals according to the North American data (Figure 3A), and then projected the unadjusted American and Australian datasets onto the layout using the same statistical model (Figure 3B). The use of unadjusted data is important here since it provides a more realistic picture of how new previously unseen samples would behave in a diagnostic pipeline. Clusters of the most frequent ALL subtypes were observable and there was a high degree of concordance between the training and validation cohorts. A visualization of all subtypes and undefined samples is available in Supplement Figures S8. For convenience, we organized the subtypes into a central supergroup (*CRLF2*, hyperdiploid, hypodiploid, *PAX5* alterations and Ph+) and distinct subtypes on the periphery (*DUX4*, *ETV6-RUNX1*, *KMT2A*, *PAX5* P80R, *TCF3-PBX1* and *ZNF384*).

**Figure 3.**
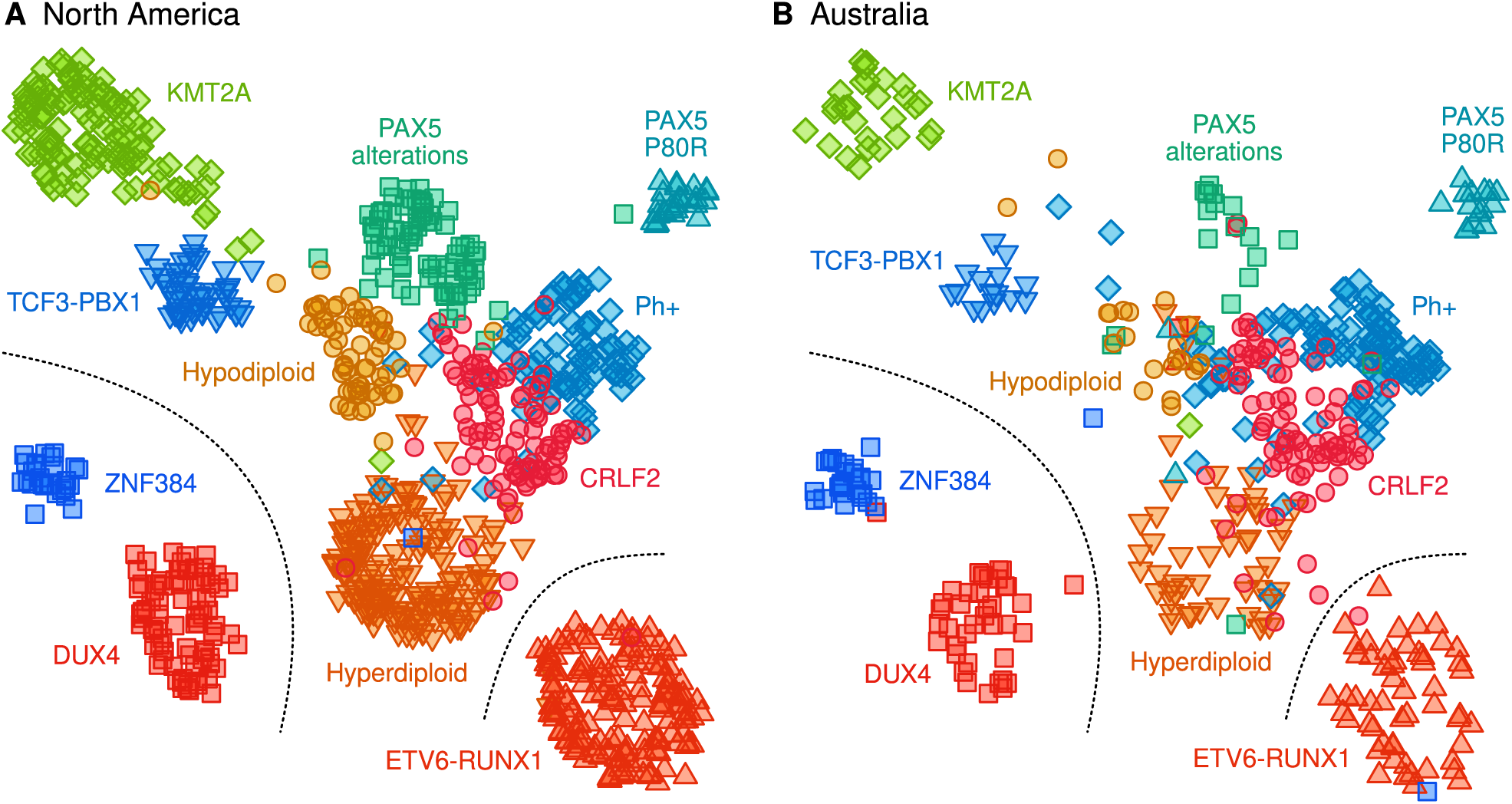
Transcriptional landscape of the most frequent ALL subtypes. Clustering structure in the North American data was modelled by the Uniform Manifold Approximation and Projection (UMAP) algorithm and using the same genes that were prioritized and pre-processed for the centroid classifier. **A**) Standardized but unadjusted gene expression values were used for re-projecting the North American samples onto the UMAP layout. We use the unadjusted expression profiles here since in practical settings where patients arrive one-by-one, adjustments for batch effects that would be available in research settings cannot be done. **B**) Unadjusted Australian data were projected onto the same UMAP layout as an independent external validation set.

### Classification of B-cell ALL subtypes based on RNA-seq profiling

Positive predictive values (PPV) of machine learning models are visualized in Figure 4 and complete performance metrics are available in Supplement Tables S1-S3. We used three different types of models (centroids, PLS and random forest, details in Methods) to exclude any artefacts that may be specific to a particular algorithm. Hyperparameters are listed in Supplement Table S4. We focused on the external validation set as the primary benchmark of accuracy. Furthermore, all performance metrics were calculated for standardized but unadjusted data to simulate a scenario where new samples are analysed one at a time without the opportunity to adjust for cohort effects.

**Figure 4.**
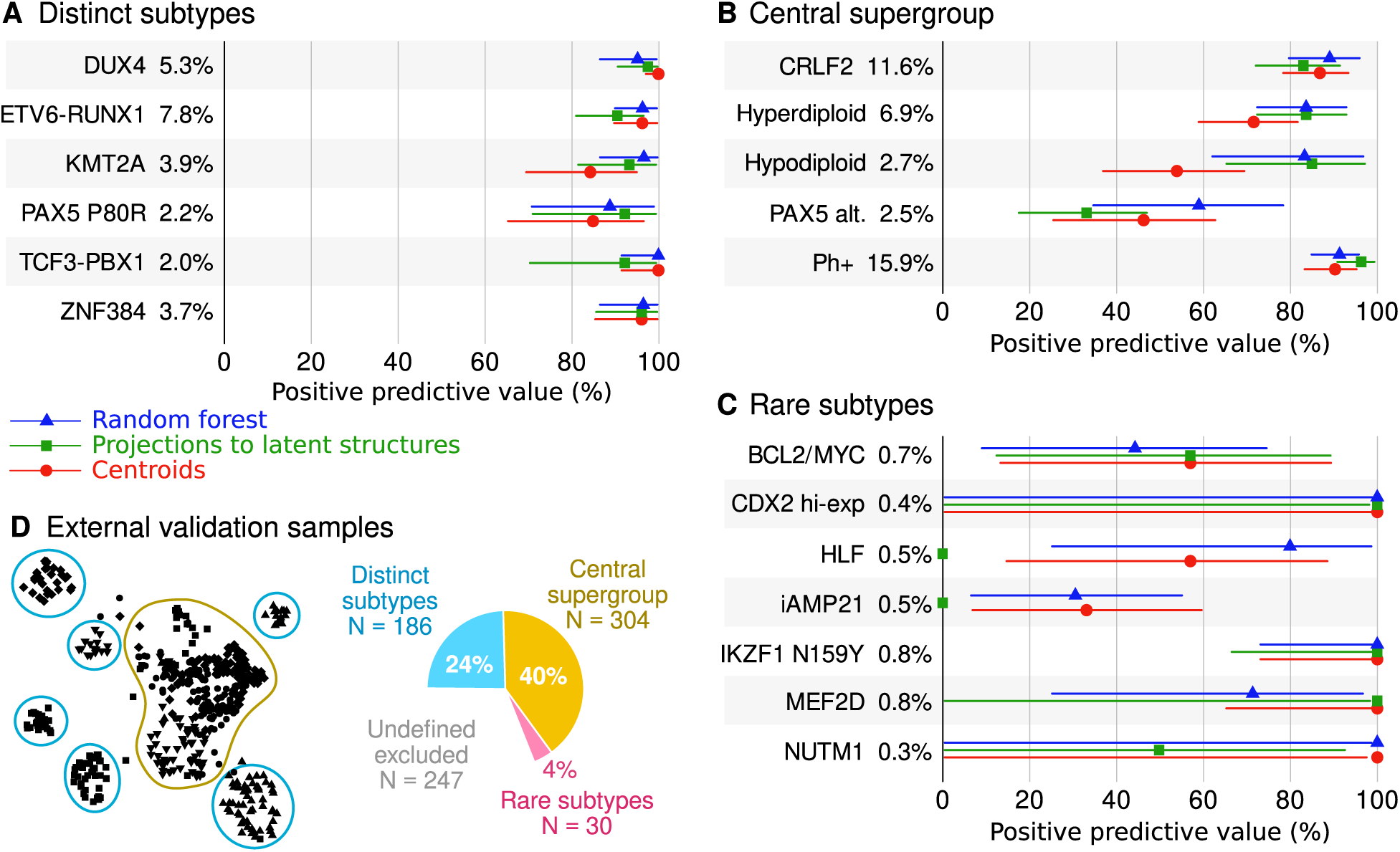
Comparison of three machine learning algorithms. Each model was fit to the batch corrected North American data (training set n = 1,078) and then evaluated in unadjusted Australian data (external validation set n = 520). Samples with undefined genetic subtypes were excluded from the analyses. The forest plots show 95% confidence intervals that reflect the statistical uncertainty due to finite category sizes. The percentages written in the plots indicate the prevalence of the genetic subtype in the Australian dataset.

Overall, differences between the three methods were negligible when considering the confidence intervals of the performance estimates (Figure 4). Accurate classification models were achieved for *DUX4* (PPV ≥ 95% in the external validation cohort across the three methods), *ETV6-RUNX1* (PPV ≥ 91%), *KMT2A* (PPV ≥ 84%), *PAX5* P80R (PPV ≥ 85%), *TCF3-PBX1* (PPV ≥ 92%) and *ZNF384* (PPV ≥ 96%). Together, 186 individuals (24%) of the Australian participants had one of these genomic subtypes (Figure 4D). More varied PPVs were observed in the central supergroup, including the *CRLF2* subtype (PPV ≥ 83%, Figure 4B), hyperdiploid (PPV ≥ 72%), hypodiploid (PPV ≥ 54%), *PAX5* alterations (PPV ≥ 33%) and Ph+ (PPV ≥ 90%). Collectively, corresponding genomic subtypes were observed in 304 individuals or 40% of Australian participants (Figure 4D). Medium to high accuracy was achieved for rare subtypes such as *IKZF1* N159Y (PPV = 100%, Figure 4C), however, the small number of cases resulted in substantial statistical uncertainty.

### Allspice classifier

Based on the results, we concluded that the centroid model of 45 prioritized genes is the preferred choice as the practical classifier due to its comparable performance to the other methods and technical simplicity. The centroids were also robust against overfitting as indicated by the flattening of internal training and testing performance when the number of inputs was increased (Supplement Figure S9). The robustness against overfitting enabled us to modify the study design to extract the maximum information from the available data (Supplement Figure S4). In the new design, the centroid model was fitted to the combined adjusted American and Australian data. We also added an extra step to account for sex and age that may carry important predictive information.

The results for a Ph+ patient are depicted in Figure 5. The classifier identifies the subtype centroid that is the most similar to the observed RNA expression profile (Figure 5A). The display includes the frequency of the subtype in the training data versus. any other subtype given the patient’s RNA profile (Figure 5B). Allspice also provides more detailed information on how the patient fits to the transcriptional landscape of ALL (technical details in Methods). The left panel shows the proximity of the patient to each subtype, respectively (Figure 5C). The *ETV6-RUNX1* subtype is an example of a genomic lesion that manifests as a clearly observable signature (Supplement Figure S10). On the other hand, there is more overlap within the central supergroup and subtypes such as hypodiploid can manifest simultaneous transcriptional proximity to multiple subtypes (Supplement Figure S11). If this overlap is too great, i.e. if it is uncertain statistically which subtype is the closest match, the patient is classified as having an ambiguous transcriptional profile (Supplement Figure S12).

**Figure 5.**
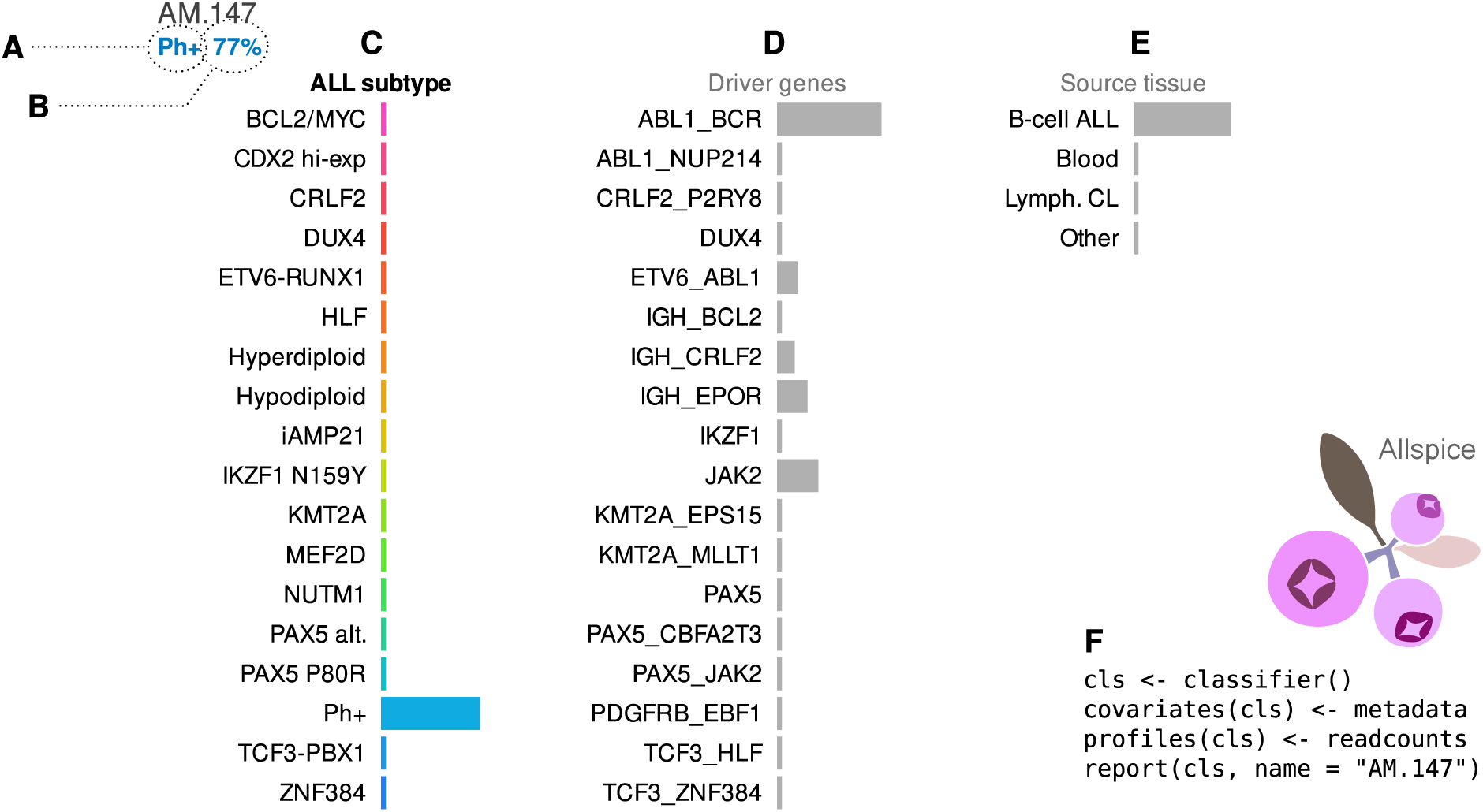
Example of a report card from the Allspice classifier for an adult male patient from North America with Ph+ genetic B-cell ALL subtype. **A**) The sample identifier and predicted subtype based on RNA data are written on the top-left corner. **B**) The report shows the frequency of the corresponding genetic subtype in the training data, given the observed gene expression profile. In this case, there is a 77% chance that sequence and cytogenetics analyses would confirm the presence of the Philadelphia chromosome. **C**) Visualization of how similar the sample is to each ALL subtype profile. The display can be interpreted as a panel of RNA “biomarkers” that are specific to each subtype. In this case, the high value for Ph+ indicates that the gene expression profile is compatible with a typical patient with the Philadelphia chromosome. **D**) Allspice also indicates how similar the sample is to the RNA profiles associated with genetic alterations. In this case, the gene expression profile matches the typical profile of patients with independently verified BCR-ABL1 fusion (i.e. the hallmark of Ph+). **E**) Samples with high leukemia burden will typically produce a strong B-cell ALL signal in the tissue panel. **F**) Minimal example of how to generate the report in the R programming environment.

The middle panel contains information on which combination of lesion-harbouring genes best fits with the observed RNA profile (Figure 5D). In this case, the gene expression profile is compatible with the classical *BCR-ABL1* fusion without other strong signals. In the release version of Allspice, we have included only mutually exclusive gene combinations with at least five cases in the training set since rarer combinations could be difficult to confirm statistically. Matching genomic alterations directly with RNA profiles may provide additional clues for samples where other diagnostic results are inconclusive (Supplement Figure S11 and S12).

The right-hand panel shows the proximity of the RNA profile to typical B-cell ALL versus other cell types from public sources (Figure 5E). For example, the Australian cohorts include patients that were recruited from routine practice, some of whom had low leukemia burden (Supplement Figure S13). For these individuals, the RNA data is closer to healthy blood and will be indicated by this panel. This feature is useful in circumstances where the sequence analyst has limited clinical information available.

### Classifier performance

The overall classification results are shown in Figure 6 and in Table 2. Of 2,046 transcriptomes, 483 (24%) were designated ambiguous and 89 (4.4%) were not classified due to poor proximity to any subtype. Performance was estimated first for all samples, including those with undefined genomic subtype. Since not all B-cell ALL cases can currently be attributed to a specific genomic lesion, these numbers are conservative estimates for the accuracy of the gene expression profile as an indicator of the underlying genomic lesion. Distinct subtypes were detected with high confidence (PPV ≥ 87%), whereas there were more ambiguous and unclassified samples in the central supergroup (see Supplement Figure S14 for a detailed break-down).

**Figure 6.**
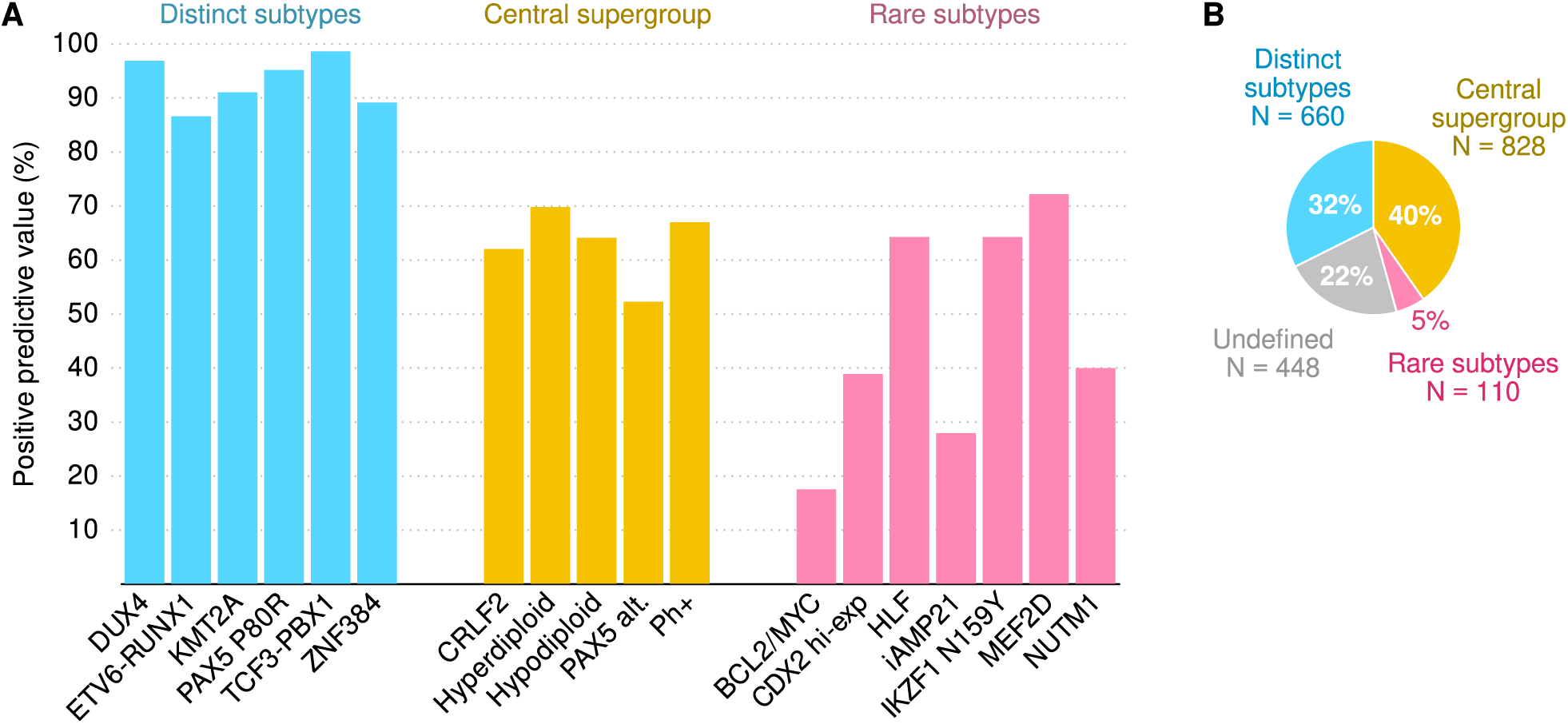
Classification performance of the Allspice centroid classifier. **A**) The bars show positive predictive values for all samples including those with undefined genetic subtypes. They represent conservative estimates on how likely it is that the subtype predicted by mRNA-seq expression levels can be confirmed as a specific sequence alteration or is also indicated by cytogenetics. **B**) Proportions of genetic subtypes in the dataset.

**Table 2.**
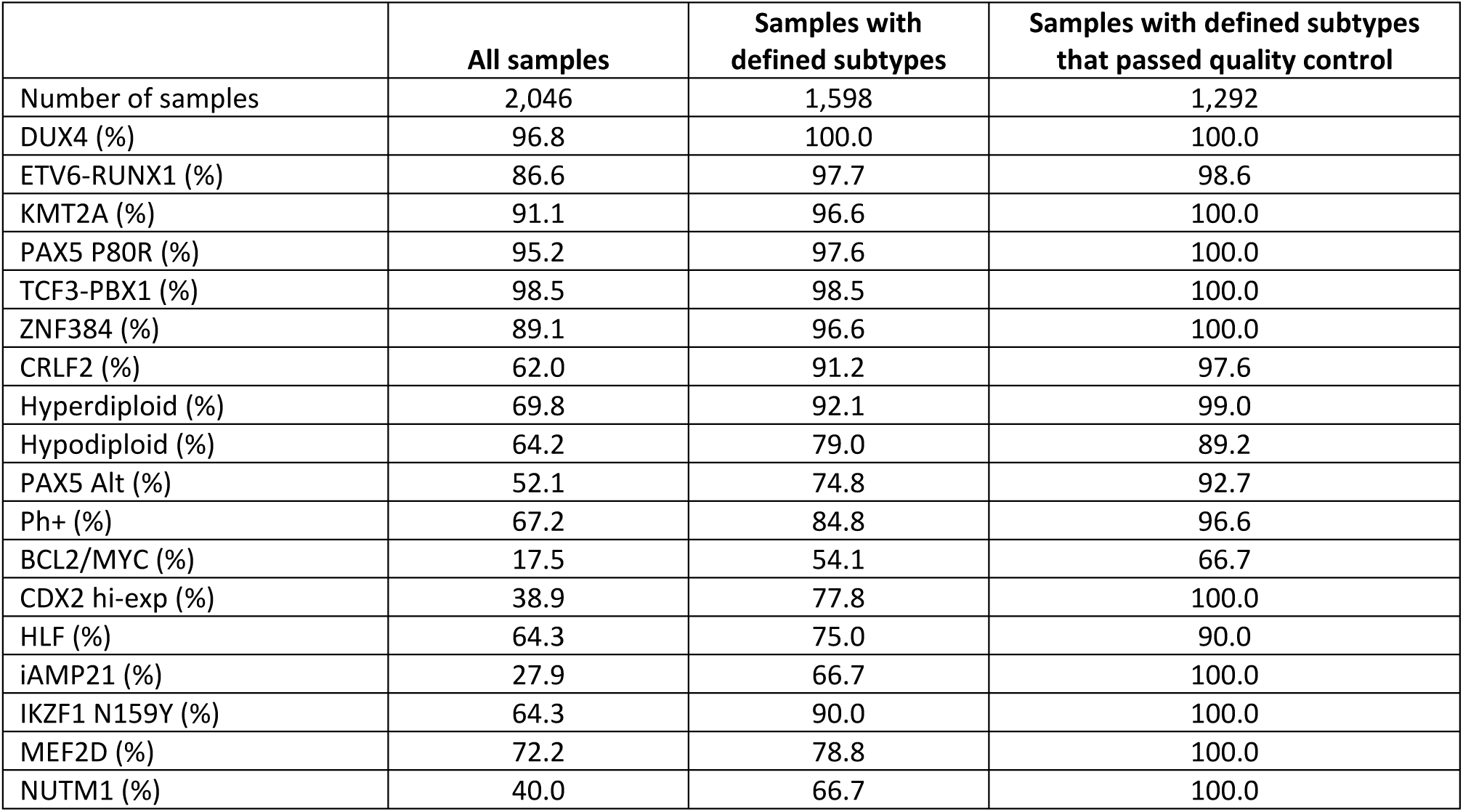
Positive predictive values for correct classification into genetically defined B-cell ALL subtypes. The values are presented as the percentages of samples for which the best matching RNA centroid was the same as the genetic subtype if defined. Quality control was set at ≥50% proximity and ≥50% exclusivity (details in Methods).

|  | **All samples** | **Samples with defined subtypes** | **Samples with defined subtypes that passed quality control** |
| --- | --- | --- | --- |
| Number of samples | 2,046 | 1,598 | 1,292 |
| DUX4 (%) | 96.8 | 100.0 | 100.0 |
| ETV6-RUNX1 (%) | 86.6 | 97.7 | 98.6 |
| KMT2A (%) | 91.1 | 96.6 | 100.0 |
| PAX5 P80R (%) | 95.2 | 97.6 | 100.0 |
| TCF3-PBX1 (%) | 98.5 | 98.5 | 100.0 |
| ZNF384 (%) | 89.1 | 96.6 | 100.0 |
| CRLF2 (%) | 62.0 | 91.2 | 97.6 |
| Hyperdiploid (%) | 69.8 | 92.1 | 99.0 |
| Hypodiploid (%) | 64.2 | 79.0 | 89.2 |
| PAX5 Alt (%) | 52.1 | 74.8 | 92.7 |
| Ph+ (%) | 67.2 | 84.8 | 96.6 |
| BCL2/MYC (%) | 17.5 | 54.1 | 66.7 |
| CDX2 hi-exp (%) | 38.9 | 77.8 | 100.0 |
| HLF (%) | 64.3 | 75.0 | 90.0 |
| iAMP21 (%) | 27.9 | 66.7 | 100.0 |
| IKZF1 N159Y (%) | 64.3 | 90.0 | 100.0 |
| MEF2D (%) | 72.2 | 78.8 | 100.0 |
| NUTM1 (%) | 40.0 | 66.7 | 100.0 |

The second set of results were calculated for 1,598 patients that had a confirmed genomic subtype. Strong performance was observed for distinct subtypes (PPV ≥ 97%) and moderate accuracy for the central supergroup (PPV ≥ 75%). This scenario captures the statistical accuracy of the classifier in ideal conditions. Thirdly, when samples that failed the proximity or exclusivity thresholds were excluded, further improvements in PPV were seen across subtypes (10 out of 18 subtypes showed PPV = 100%). This shows how assessing the sample quality will help to avoid misclassification of borderline cases. Of note, a high proportion of samples (56%) that were classified as having *BCL2/MYC* gene expression subtype were also identified as not originating from ALL B-cells in the tissue classifier (Supplement Figure S15) which may explain why it was the most difficult subtype to predict.

## Discussion

B-cell ALL remains a life-threating disease particularly for adult patients of specific genomic subtypes [1–3,9]. Recently, rapid progress has been made in detecting ALL subtypes by RNA sequencing [12–15,19] and in subtype-specific treatments [6, 20]. In this study, we present new data from a large Australian dataset and new findings from rigorous statistical and practical considerations to better leverage gene expression profiling in the diagnosis of ALL subtypes. We confirmed six genomically defined subtypes in a third of patients that produce highly predictable mRNA profiles (PPV ≥ 89%). Further 40% of individuals were distinguishable by mRNA-seq expression levels, although the associations between specific genomic lesions and transcriptomic profiles were less certain. To dissect the biological ambiguity, we developed a proof-of-concept classifier that aggregates genome-wide mRNA-seq read counts into simplified RNA biomarker scores that indicate how well and where a patient’s RNA profile fits in the landscape of B-cell ALL subtypes.

### Definition of ALL subtypes

We used simpler definitions of genomic ALL subtypes compared to some of the previous reports [12, 21]. The streamlined presentation provided multiple benefits although trade-offs were unavoidable. Firstly, it allowed for sufficient group sizes for robust statistics and a statistically meaningful overview of the transcriptional landscape (e.g. Figure 3). On the other hand, more granular information on the exact nature of sequence alterations may be of high clinical importance but not captured by our subtype definitions. To gain better mechanistic insight, the Allspice classifier includes a feature that indicates genes that may harbour a driver mutation (Figure 5D) and further development of this concept may enable accurate diagnostics for targetable gene expression abnormalities. We also designed Allspice to support subtype definitions that are not mutually exclusive, thus providing flexibility for future updates.

The second rationale for the streamlined ALL taxonomy was to ensure a rigorous study design for machine learning. Previous studies have classified patients according to the way their RNA-seq expression profiles cluster (examples include the Ph-like and *ETV6-RUNX1*-like subtypes [12, 13]). However, these definitions are problematic for the training of RNA profiling classifiers – the same gene expression levels should not be used to first define and then predict a subtype. Instead, we relied on observable sequence abnormalities or other information that was not derived from gene expression levels (except for *DUX4*). As a trade-off, the size of the training set decreased and the number of individuals with undefined genomic subtypes increased, however, such uncertainty is to be expected in real-world datasets that manifest substantial biological variation in how genomic lesions drive altered gene expression profiles.

### Classification performance and utility

Overall, B-cell ALL subtypes that could be identified by fusion callers and cytogenetics had distinctive mRNA-seq read count profiles (Allspice classified 90% of samples with a defined genomic subtype correctly). In a recent study that used mostly the same datasets, correct classification rate was between 82% and 93% [21] and similar rates have been reported in other machine learning studies of ALL [11,19,28–30]. Therefore, the performance of the Allspice tool is within the range of other similar classifiers, which demonstrates the rich biological information available from RNA-seq data and the stability of the predictions across multiple types and implementations of classifiers.

If a patient tests positive by Allspice for one of the six distinct subtypes (*DUX4*, *ETV6-RUNX1*, *KMT2A*, *PAX5* P80R, *TCF3-PBX1* and *ZNF384*), our findings suggest that the subtype can be validated by deeper exploration of the sequencing reads in the same RNA-seq dataset or by molecular diagnostics for at least 89% and up to 99% of cases depending on the subtype. Both the sequencing and molecular analyses can be time-consuming and inconclusive, whereas mRNA expression levels (i.e. inputs to Allspice) can be reproducibly calculated using highly standardized algorithms. This will shorten the time to diagnosis for the vast majority of ALL cases with the aforementioned recurrent lesions, and significantly shorten the time to delivering care in the clinical setting. Identification of other lesions such as *CRLF2*, hyperdiploid, hypodiploid, *PAX5* alterations and Ph+ are less definitive, though this may improve with further algorithm training and refinement. There may be clinical utility for the Allspice biomarker panels as RNA risk factors for adverse outcomes in patients that show abnormal karyotypes.

A total of 448 of patients lacked a definitive genomic subtype under the streamlined taxonomy we used in this study. Diagnostics for this patient subpopulation is where we expect mRNA-seq profiling to provide the best added value. In this respect, Allspice is a unique tool since it also provides quantitative RNA biomarkers for the most likely driver lesions and for the deviation from the healthy blood transcriptome. These features are particularly useful when the ALL subtype is difficult to establish due to the absence of identifiable sequence alterations or inconclusive cytogenetic findings. Based on the RNA data, we assigned a transcriptionally compatible ALL subtype to 207 patients (41% of 448) – these are conceptually similar to the Ph-like and *ETV6-RUNX1*-like gene expression profiles from previous studies. The same concept can be extended to genomic drivers. For example, there were 90 (20%) patients with simultaneous alterations in CRLF2 and P2RY8 and 79 (18%) patients with alterations in both IGH and BCL2 among the undefined subpopulation. Given the sizes of these secondary subgroups, they may be considered as additions or replacements for historical subtypes as genomic ALL datasets get larger and better phenotyped. The gene expression profiles of 57 (13%) patients could not be matched with any typical ALL subtype and we suspect many of these were individuals with a low leukemia burden.

### Strengths and weaknesses

The large sample size (statistical power), diversity of data sources, careful mitigation of potential confounding factors and comparisons between three classes of machine learning algorithms make this study strong from a methodological perspective. Notably, the Australian samples were collected from routine health care settings, which provides a realistic spread of sample quality and leukemia burden as encountered in clinical practice. Furthermore, we used additional datasets to help assess the quality of the samples and safety against misclassification, which is an important practical consideration outside research settings [31]. The data were obtained from three Western countries, and caution is warranted if applying the findings in different ethnic or socioeconomic context. Due to the careful analyses and robust performance, we anticipate that the classifier we created captured biological information that reflects the causal mechanisms of ALL, and it is therefore likely to work well for most patient populations.

### Practical considerations

Allspice is open source, easy to install on the popular R programming environment via the Comprehensive R Archive Network and it comes with extensive documentation. It accepts raw read count data as produced by standard RNA sequencing pipelines, which is an advantage in clinical settings that may lack a dedicated bioinformatician. The R environment already includes tools for visual clustering of transcriptomes using algorithms such as t-SNE [32], but clustering results can be difficult to interpret for individual cases. Rather than relying on visual proximity in a scatter plot, Allspice uses quantitative probabilistic metrics to indicate the certainty of the predicted subtype. Furthermore, the ability to analyse one sample at a time is important: t-SNE or hierarchical cluster analysis are designed for the research space where large cohorts of labelled samples are readily available, whereas Allspice was designed for a single sample from the beginning.

We included two examples with unusual lesions where Allspice helped to assign a (transcriptional) subtype. The first case was an individual with an undefined genomic subtype that Allspice classified as having an RNA profile compatible with an ETV6-RUNX1 fusion. Uncommon ETV6 fusions were discovered by detailed investigations (Supplement Figure S16). Another individual was classified as having a ZNF384-like transcriptional profile while the exact causal lesion remained uncertain (Supplement Figure S17). These examples highlight how the additional information from Allspice can guide diagnostic efforts for patients with unusual genomic lesions.

## Conclusions

We observed strong associations between genomic alterations and lymphoblast transcriptome-wide expression profiles in paediatric and adult patients of B-cell ALL. For a third of patients, these associations are unambiguous and provide diagnostic information that is often quicker and easier to obtain compared to fusion callers or cytogenetic tests. For the rest of the patients, gene expression analyses may provide insight that is not available from other methods. An RNA-based ALL biomarker can inform sequence analysts where to look for lesions manually if automatic fusion callers failed or read counts were too low for statistical certainty. In both scenarios, Allspice can help oncologists to determine the most likely causal drivers with greater confidence and identify potential therapeutic targets in a shorter time frame.

## Methods

### North American participants

A total of 649 males, 541 females and 89 patients without gender information were from St Jude Children’s Research Hospital (St Jude); Children’s Oncology Group (COG); ECOG-AGRIN Cancer Research Group (ECOG-AGRIN); MD Anderson Cancer Center (MDACC); the Alliance of Clinical Trials in Oncology, Cancer and Leukemia Group B (CALGB) and University of Toronto (Toronto). Detailed clinical information for each case and listings of clinical trial numbers have been previously published [12]. RNA-seq data files were obtained from the European Genome-Phenome Archive (EGAD00001004461 and EGAD00001004463). The patients who participated in this study have provided written informed consent, assent (as appropriate), or parental consent (as appropriate) as part of enrolment protocols, for research, including genetic research. All relevant ethical regulations were followed during this study.

### Australian participants

A total of 387 males, 278 females and 102 patients without gender information were investigated through the Australasian Leukaemia and Lymphoma Group (ALLG) National Blood Cancer Registry and the associated Regalia project (ACTRN12612000337875), Australian & New Zealand Children’s Haematology/Oncology Group Acute Lymphoblastic Leukaemia Study 8 (ACTRN12607000302459) and Study 9 (ACTRN12611001233910). All protocols had been approved by relevant human research ethics committees.

### Supporting RNA data

RNA data were sourced from a previously published study of 660 lymphoblast cell lines [33] and from the Genotype-Tissue Expression (GTEx) project release 8 [34]. The GTEx includes RNA-seq data from 948 donors and 54 tissues. In this study, we organized the data into 31 organ groups, of which those that contained at least 500 samples were selected, including whole blood as the most relevant tissue type for ALL.

### RNA sequencing and pre-processing

RNA analyses of the North American samples have been described previously [12]. Briefly, RNA-seq was performed using TruSeq library preparation and HiSeq 2000 and 2500 sequencers (Illumina Inc, San Diego, CA, USA). All sequence reads were paired-end and were obtained from total RNA and stranded RNA-seq (75 or 100 base-pair reads) and polyA-selected mRNA (50, 75 or 100 base-pair reads). In Australia, library preparation for mRNA sequencing was performed using either Truseq Stranded mRNA LT Kit (Illumina, CA, USA) or Universal Plus mRNA-Seq with NuQuant (Tecan, CA, USA) from 1 g of total RNA as per manufacturer’s instructions. Samples were sequenced by either Illumina HiSeq 2000 or NextSeq 500 platforms producing 75b length paired-end (PE) reads with median read depth of 65M reads.

Raw reads from all cohorts were aligned and mapped to the GRCh37 reference genome with the STAR software version 2.4.2a and above using the two-pass mode [35]. Raw gene counts were generated from BAM files using featureCounts [36]. We defined a gene to be usable as a potential biomarker if it had a read count of ≥100 in at least 1% of samples in both the North American and Australian datasets, respectively. In total, 18,923 genes were included in the study. Expression counts were normalized using the DESeq2 algorithm [37] and the normalized counts were transformed using the formula *log_2_(count + 1)* before statistical analyses.

### Genomic subtyping

In the text, we use the term ‘genomic ALL subtype’ when the subtype is assigned according to a DNA or RNA sequence abnormality or a clinical biomarker independently of gene expression levels. The detection of genomic alterations and subsequent subtyping of ALL cases were based on the previously published analyses of the North American samples [12] and a preliminary definition of a recently discovered rare CDX2 subtype [38]. We were not able to define the DUX4 subtype independently of mRNA expression levels due to its cryptic nature.

Genomic alterations in the Australian samples were detected as follows. FusionCatcher [39], SOAPfuse [40] and JAFFA [41] were utilised to identify clinically relevant gene fusions. Only fusions reported by multiple fusion calling algorithms were considered with the exception of rearrangements involving the IGH locus (IGH-DUX4), which were confirmed by high levels of DUX4 expression [42] or by manual inspection of the fusion and accompanying expression data. Single nucleotide variants were identified with GATK-haplotype caller [43] following the best practices workflow. Copy number alterations were detected by multiplex ligation-dependent probe amplification using two SALSA Reference kits (P335 and P202, MRC-Holland, Amsterdam, Netherlands) according to the manufacturer’s instructions.

To harmonize subtype definitions between the cohorts and to include only the most confident classifications, we relabelled subtypes for the statistical analyses (Supplement Table S1, Supplement Figure S1). In addition to the pre-defined subtypes, we also determined every gene and combinations of genes that harboured an alteration for any individual. These mutational profiles were then used for creating genetically matched subsets to allow for more accurate batch adjustments (details below).

### Adjustments for confounders

We divided the data into five batches according to country of origin and age of the patients (Table 1). We then used statistical adjustments to mitigate the potentially confounding associations between data sources and ALL subtypes. Firstly, we used Surrogate Variable Analysis [44] to reduce undesired variation in normalized read counts within each batch, respectively. Next, we used genetically matched subsets and the Numero R package [45] to remove potentially confounding variation between the batches (Supplement Figure S2). Genes that were perfectly aligned with a batch were excluded, which left 18,503 adjusted genes (98%) for statistical analyses. We then calculated correlations between unadjusted and adjusted versions of gene expression and excluded unstable genes that showed a Pearson correlation of R < 0.9 (Supplement Figure S3). A total of 6,673 genes were considered stable and included as inputs to classification models.

### Machine learning

We chose a random forest approach [46] as an example of a supervised non-linear machine learning technique that can predict multi-class outcomes from complex input data. We also created Projections to Latent Structures (PLS) for each ALL subtype separately as an example of a linear factorization method [47]. Separate PLS models were fitted to each subtype versus other samples. The third type of modelling was based on neighbour distances: we calculated mean RNA profiles (centroids) for every ALL subtype and classified individuals based on the nearest centroid in data space. Unsupervised clustering was achieved with the Uniform Manifold Approximation and Projection (UMAP) algorithm [48] to gain qualitative visual insight into the RNA-based segregation of ALL subtypes.

Input data were standardized the same way for each model using the default settings of the function ‘numero.prepare()’ in the Numero R package [45]. This is a refined version of the empirical Z-score with protections in place for outliers and skewed distributions. Normalization and standardization parameters were determined for the training dataset and applied independently to the external validation set to simulate a scenario where new unseen data are analysed one sample at a time and thus must be pre-processed using pre-defined parameters.

### Pruning of correlated input features

To improve the performance of UMAP and nearest centroids, the full list of detectable genes was pruned using an approach similar to clumping in genetics [49] and the pruned set of genes was used as input features. First, we calculated Welch’s t-statistics for each gene and ALL subtype and converted them to Z-scores using the cumulative t-distribution and inverse cumulative Normal distribution. Next, we calculated the variance of the Z-scores for each gene as an aggregate measure of how well a gene segregated between ALL subtypes. Genes were then sorted from large to small variance. In the final step, the sorted list was traversed while checking if the next gene was correlated (R ≤ −0.3 or R ≥ 0.3) with any of the already selected. The Australian data were excluded from the pruning procedure to ensure independent external validation.

### Training, testing, external validation and performance metrics

The division of data for the evaluation of classification performance is shown in Figure 1. To determine the optimal model complexity (hyperparameters), North American participants were divided into two randomized subsets that were used as internal training and testing sets. The randomization was done separately for each subtype to ensure matching subtype frequencies. Next, models were fitted to one of the subsets (internal training set) and classification performance was evaluated in the other (internal testing set). Classification performance in the testing set was used to determine optimal hyperparameter settings (Figure 1E).

Full models were trained with the full North American dataset and validated externally in the Australian dataset. We focused on positive predictive value (PPV) as the primary performance metric due to its suitability for the low case frequencies of most ALL subtypes. Negative predictive values, sensitivity, specificity and the area under the receiver operating characteristic curve were also calculated.

### Proximity and exclusivity

The open-source classification tool Allspice is available in the Comprehensive R Archive network (URL: https://cran.r-project.org). It includes a visual display of the classification results (Supplement Figure S4, Box F) and two quality measures to help decide if an RNA profile indicates a specific ALL subtype. The proximity measure is the output value from the probit regression step in the Allspice modelling design (Supplement Figure S4, Box E) and it represents the likelihood of observing the subtype in the training population when balanced for group sizes and given the observed RNA biomarker value and covariates. We chose 50% as the threshold for acceptable proximity. To identify samples with mixed subtype characteristics (designated as ‘Ambiguous’), we defined exclusivity as the difference in the proximity scores between the best and the second-best matching subtype centroids. The χ^2^-distribution with one degree of freedom was used to convert the difference into a probability. We chose 50% as the default threshold for acceptable exclusivity.

## Data Availability

The North American mRNA-seq data files are available in the European Genome-Phenome Archive (EGAD00001004461 and EGAD00001004463). The Australian dataset is available from the authors upon request.

https://ega-archive.org/datasets/EGAD00001004461

https://ega-archive.org/datasets/EGAD00001004463

https://gtexportal.org/home/datasets

https://www.ebi.ac.uk/arrayexpress/experiments/E-GEUV-1/files/analysis_results/

## Declarations

### Ethics approval and consent to participate

North American patients who participated in this study have provided written informed consent, assent (as appropriate), or parental consent (as appropriate) as part of enrolment protocols, for research, including genetic research [12]. Australian ethics approvals were obtained as part of the Australasian Leukaemia and Lymphoma Group (ALLG) National Blood Cancer Registry and the associated Regalia project (ACTRN12612000337875), Australian & New Zealand Children’s Haematology/Oncology Group Acute Lymphoblastic Leukaemia Study 8 (ACTRN12607000302459) and Study 9 (ACTRN12611001233910).

### Consent for publication

Individual consent for publishing the RNA profiles of the Australian patient CHI_0809 was obtained by the Children’s Cancer Institute and patient CHII_0804 by the Australian Genomics Health Alliance.

### Availability of data and materials

The Allspice software and source code are available via the Comprehensive R Archive Network (URL: https://cran.r-project.org). The North American mRNA-seq data files are available in the European Genome-Phenome Archive (EGAD00001004461 and EGAD00001004463). The Australian dataset is available from the authors upon request.

### Authors’ contributions

VPM developed the machine learning models and analyzed RNA read counts, JR determined genomic ALL subtypes, JR and JB obtained and processed raw mRNA-seq data, DY and DLW contributed to the study design. All authors edited and reviewed the manuscript.

## Acknowledgement

We are grateful for the contributions by Australian leukemia patients and health care professionals for providing us with the original biological material, including the Australian and New Zealand Children’s Haematology/Oncology Group. We also acknowledge the assistance of the South Australian Genomics Centre for generating and analysing RNA-seq data.

---

**Supplement Figure S1.**
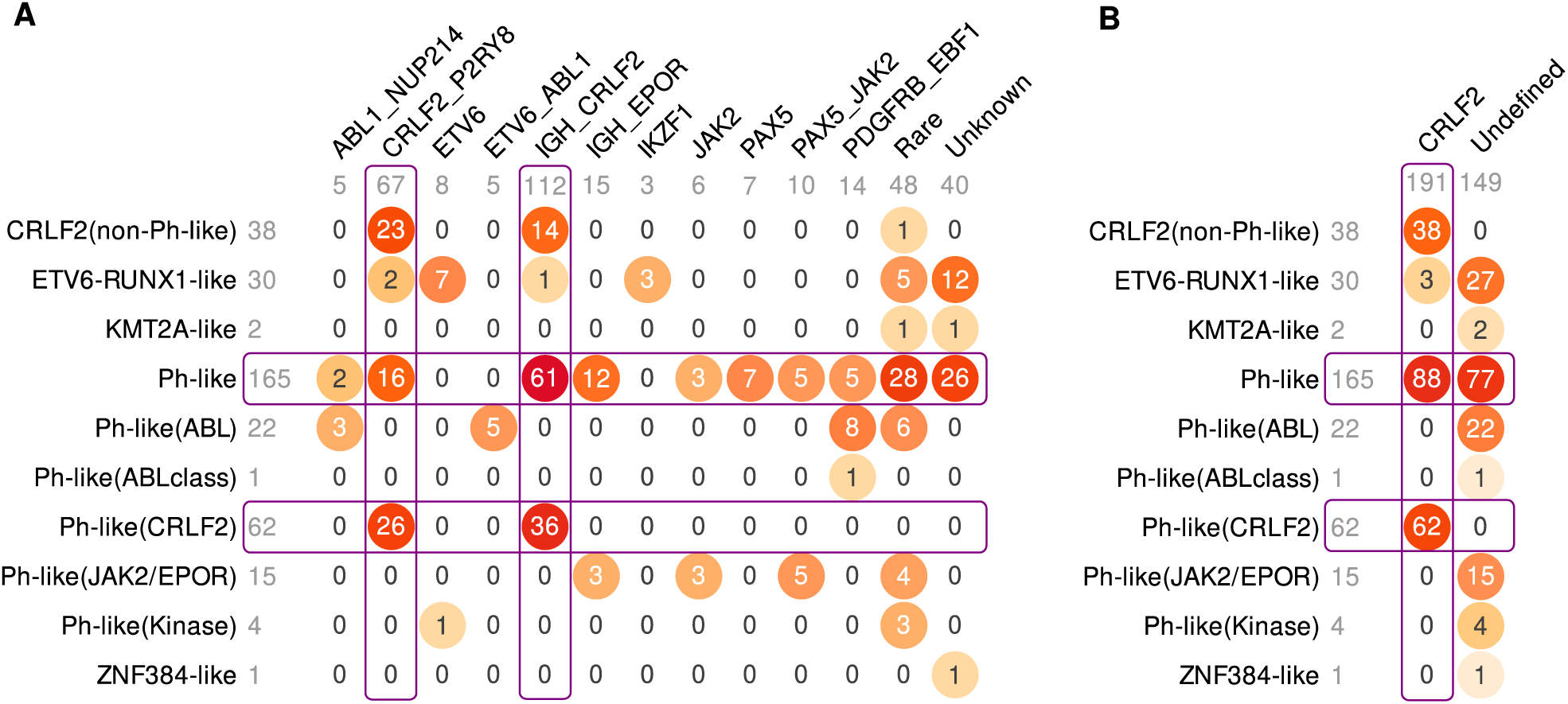
Re-definition of subtypes that were derived predominantly from clustering of gene expression data. From a machine learning perspective, it would be problematic to first define a subtype using gene expression profiles, and then predict the subtype using the same data (circular design). To preserve sound design for this study, the Ph-like subtype was not used as the target for prediction by RNA-seq data. **A**) Overlap between RNA-based “like” subtypes (vertical) and combinations of gene alterations (horizontal). **B**) Patients that were Ph-like or otherwise derived from RNA-seq clustering were assigned to the CRLF2 subtype if they harboured alterations involving the CRLF2 gene as this was the most frequent genetic lesion within these patients.

**Supplement Figure S2.**
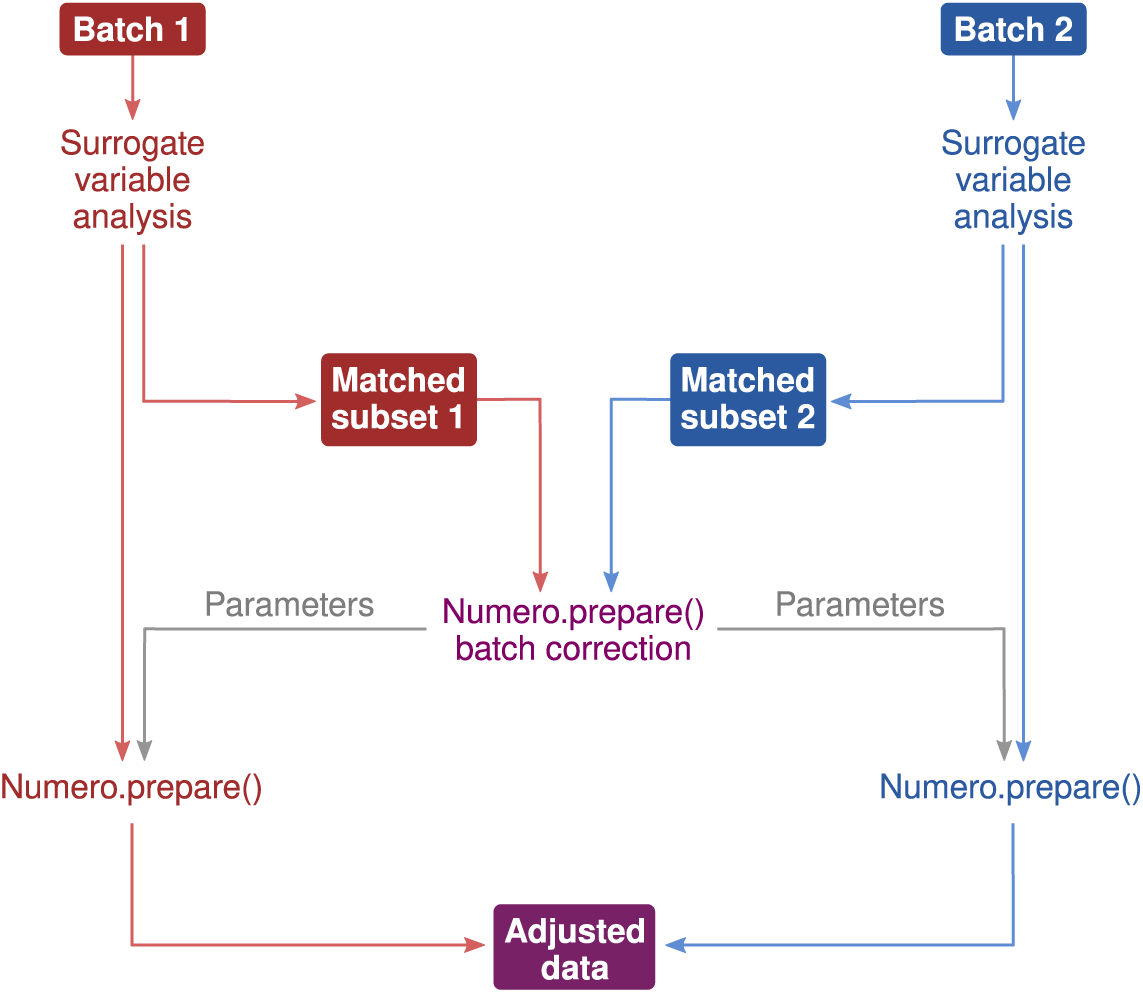
Schematic illustration of batch corrections. The RNA-seq datasets contained undesirable correlations between gene expression and multiple potential confounders including library strandedness, subtype prevalence across different locations and other cohort effects. To prevent the classification models from fitting to these patterns, we first applied surrogate variable analysis (SVA) to remove variation that was not related to the traits of interest within each batch. Note that applying SVA to the entire dataset could lead to over-optimistic classifiers, since it would amplify existing correlations between batch membership and the subtypes that arise from the cohort structure. To remove the correlations between batch membership and biological subtypes, we created matched subsets from each batch that had pair-wise identical age, sex, genetic subtype and known lesion profile. It thus became safe to do standard batch correction using the Numero library for these subsets. Lastly, the adjustment parameters from the subset analysis were used for adjusting the batch effects of the original data.

**Supplement Figure S3.**
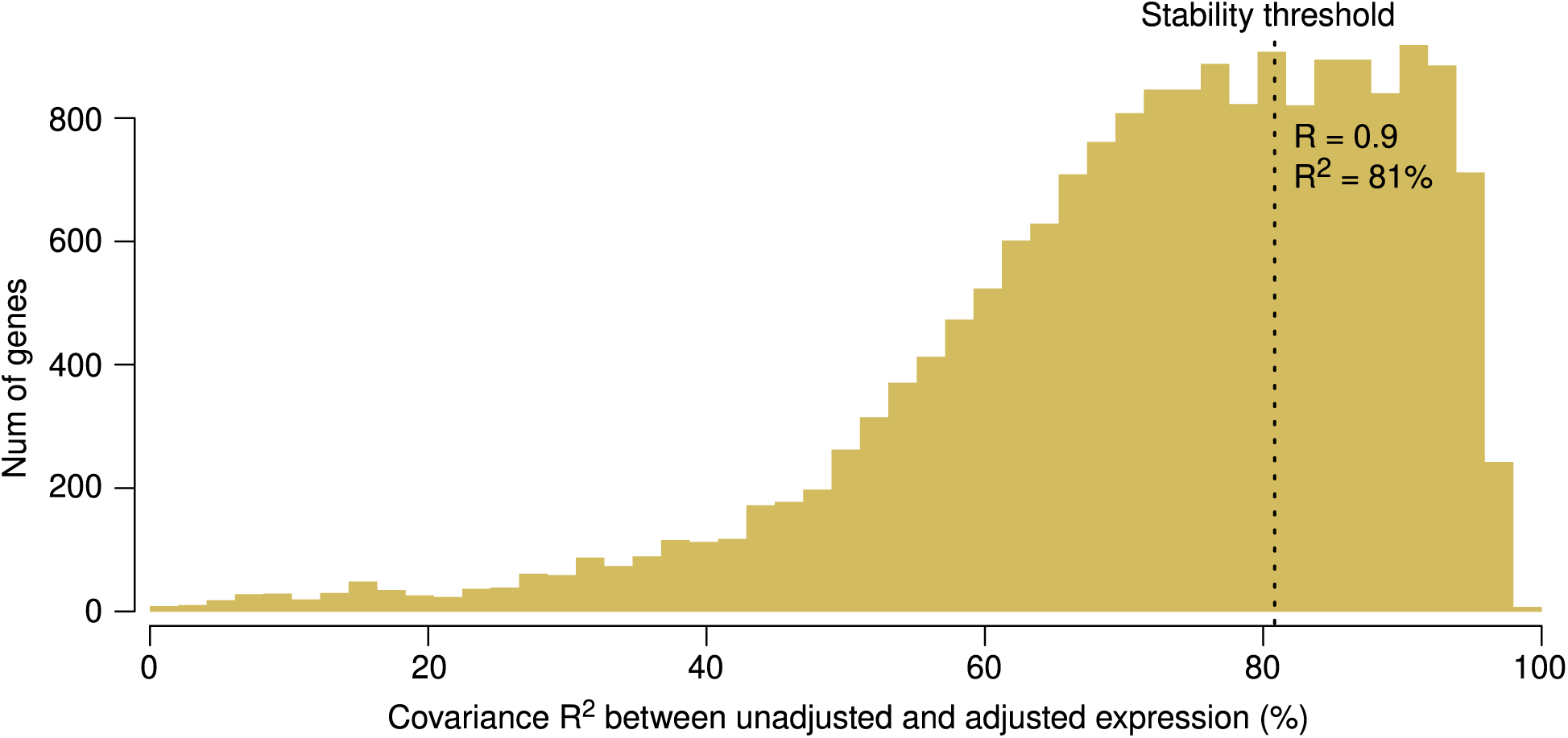
A summary of the impact of batch correction on gene expression levels. The histogram shows Pearson correlations for each gene between original log transformed read counts and read counts after batch correction. We then defined acceptable stability as R > 0.9, which led to the inclusion of 6,249 genes out of 18,515 for further statistical analyses.

**Supplement Figure S4.**
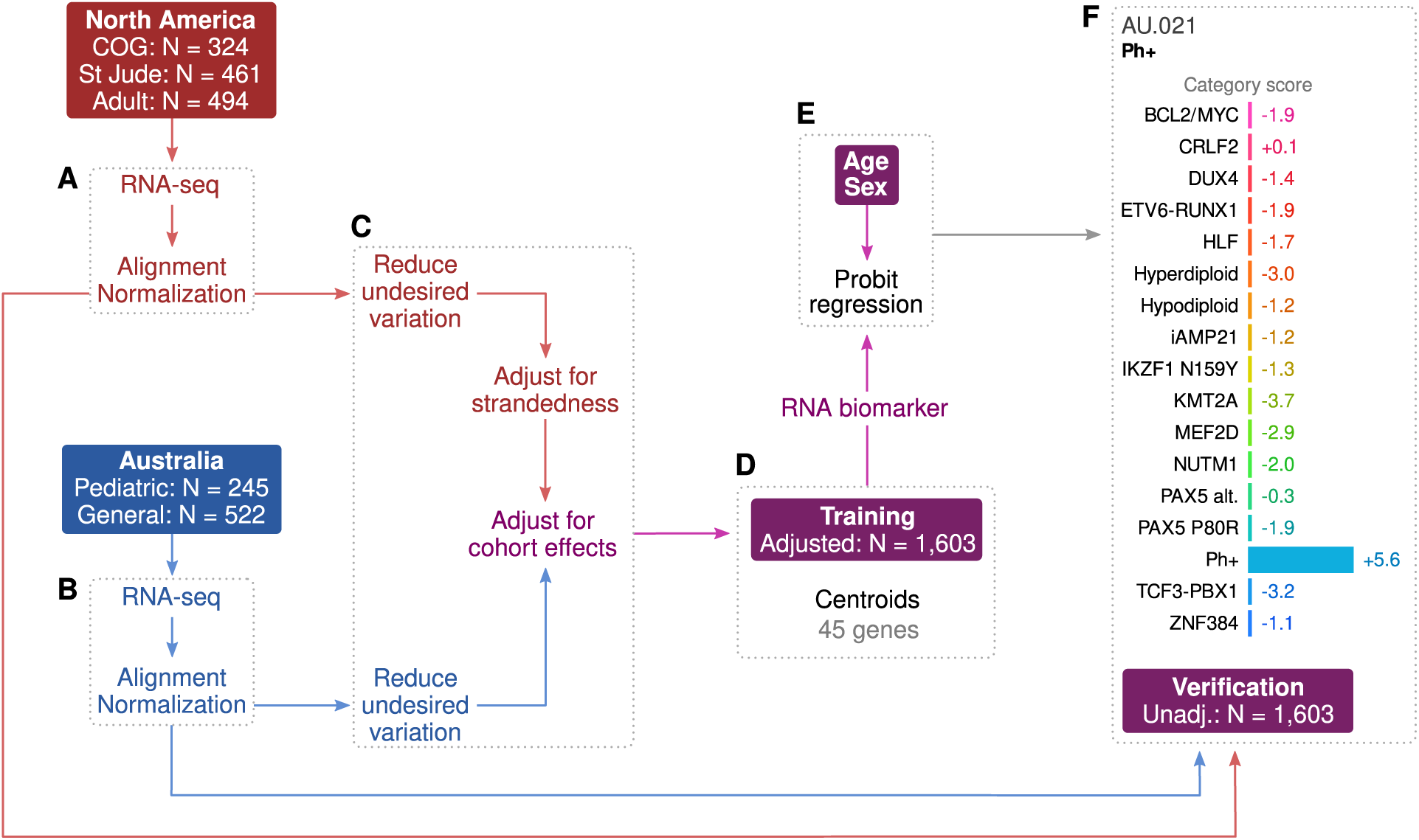
Centroid classifier as trained for the Allspice R library. All available samples that had a verified genetic subtype were used as a training set. Altogether 45 inputs out of 6,673 stable genes were prioritized according to the clumping algorithm described in Methods. The model was trained with batch corrected data and performance metrics were calculated from unadjusted expression levels.

**Supplement Figure S5.**
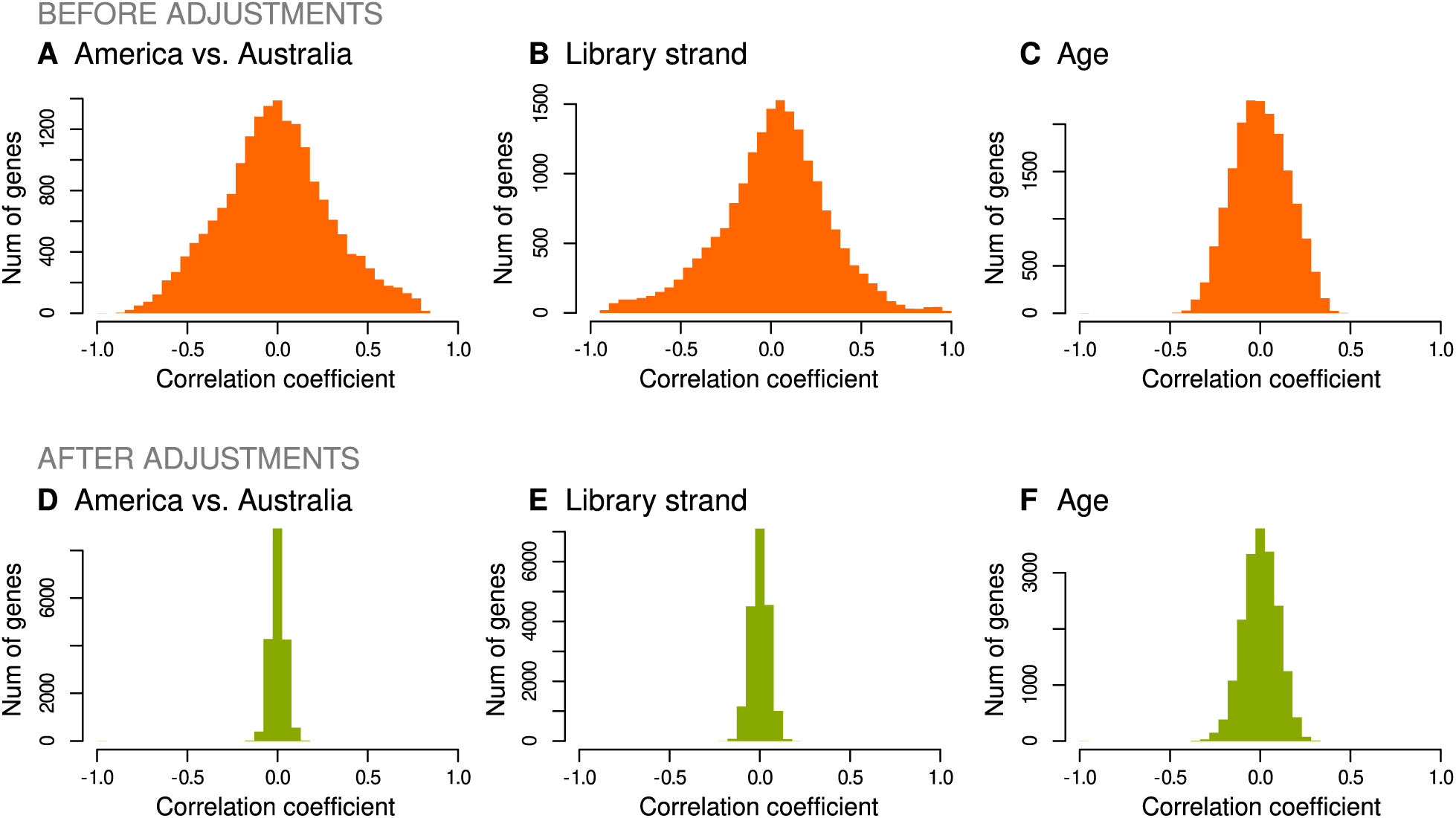
Comparison of Pearson correlations between log-transformed read counts and patient meta-data before and after batch correction. Age was not used as a matching criterion in those batch correction steps that involved comparisons between pediatric and non-pediatric batches.

**Supplement Figure S6.**
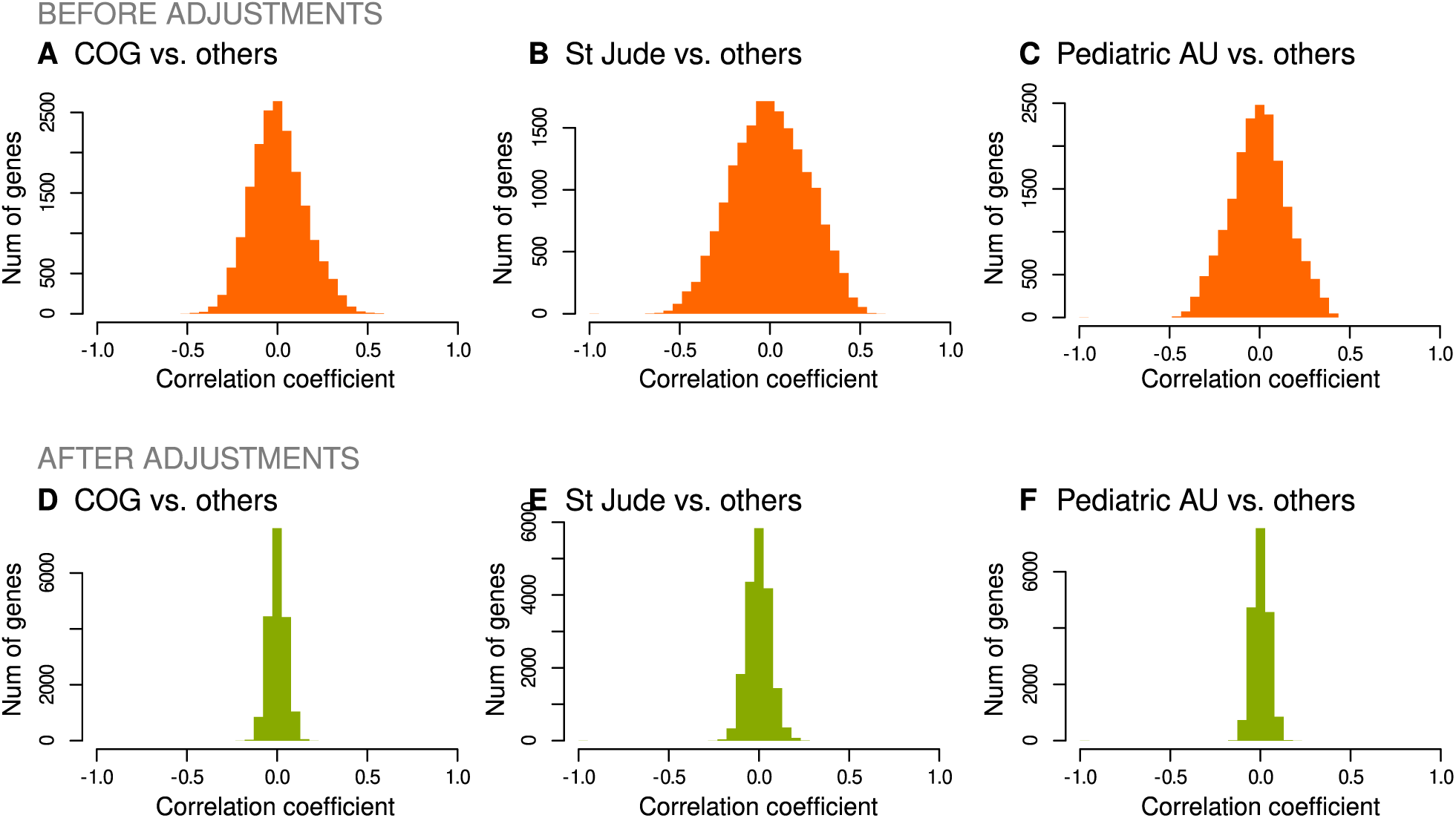
Comparison of Pearson correlations between log-transformed read counts and patient meta-data before and after batch correction. Age was not used as a matching criterion in those batch correction steps that involved comparisons between pediatric and non-pediatric batches.

**Supplement Figure S7.**
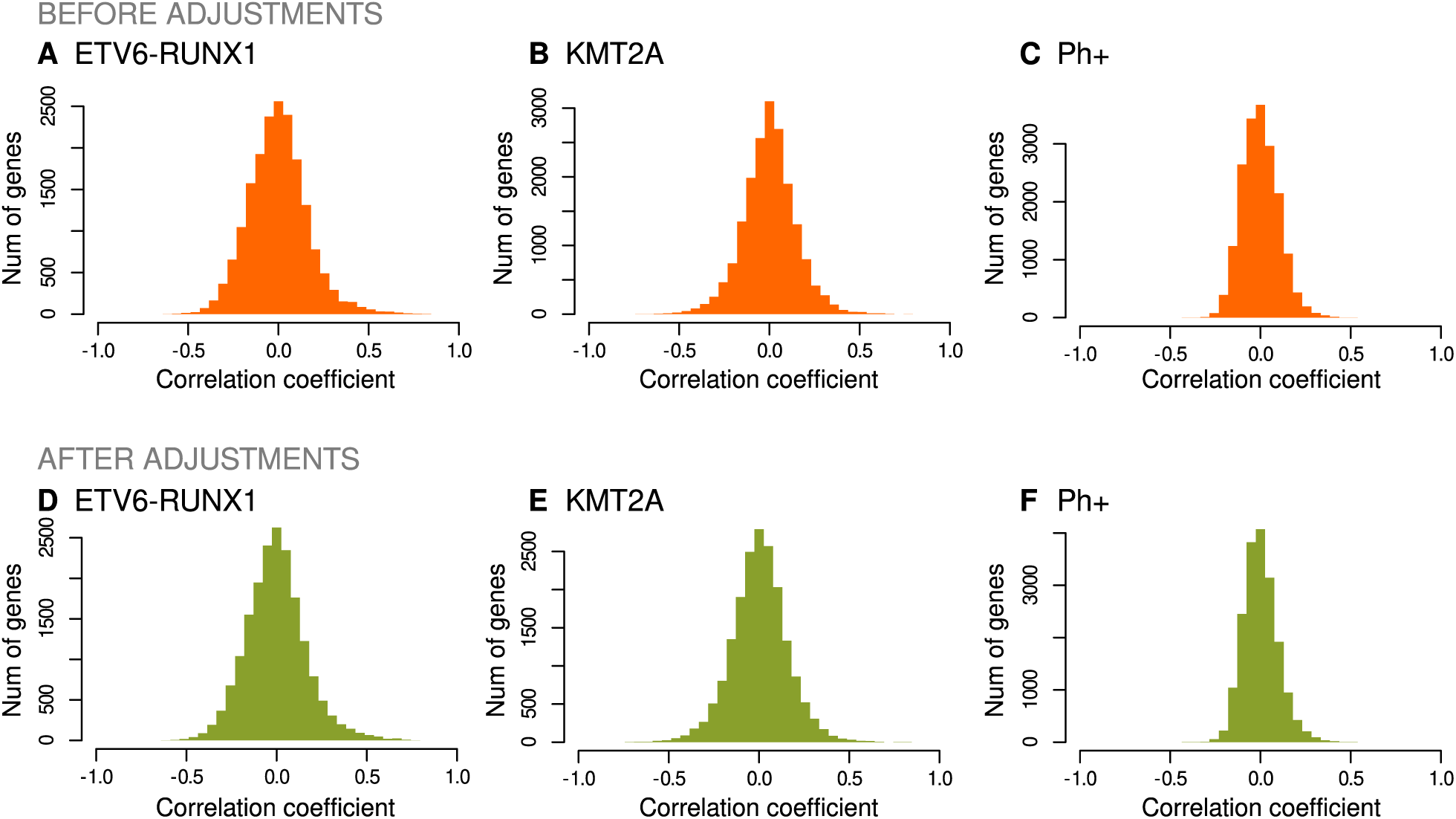
Comparison of Pearson correlations between log-transformed read counts and patient meta-data before and after batch correction.

**Supplement Figure S8.**
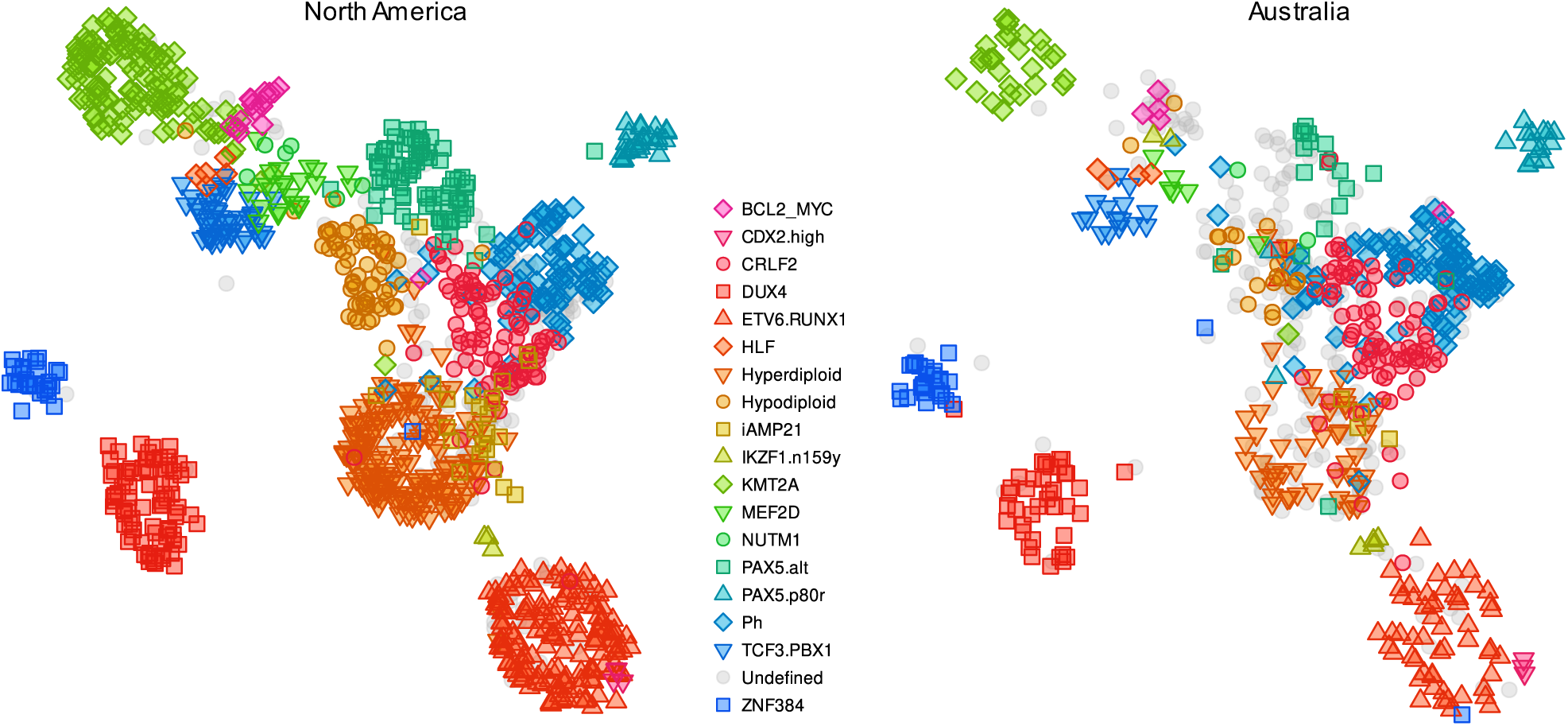
Uniform Manifold Approximation and Projection (UMAP) trained with batch corrected North American data and using genes that were optimised the centroid classifier. The scatter plot was produced by applying the UMAP to the unadjusted North American dataset and Australian datasets, respectively. The grey symbols represent undefined genetic subtypes.

**Supplement Figure S9.**
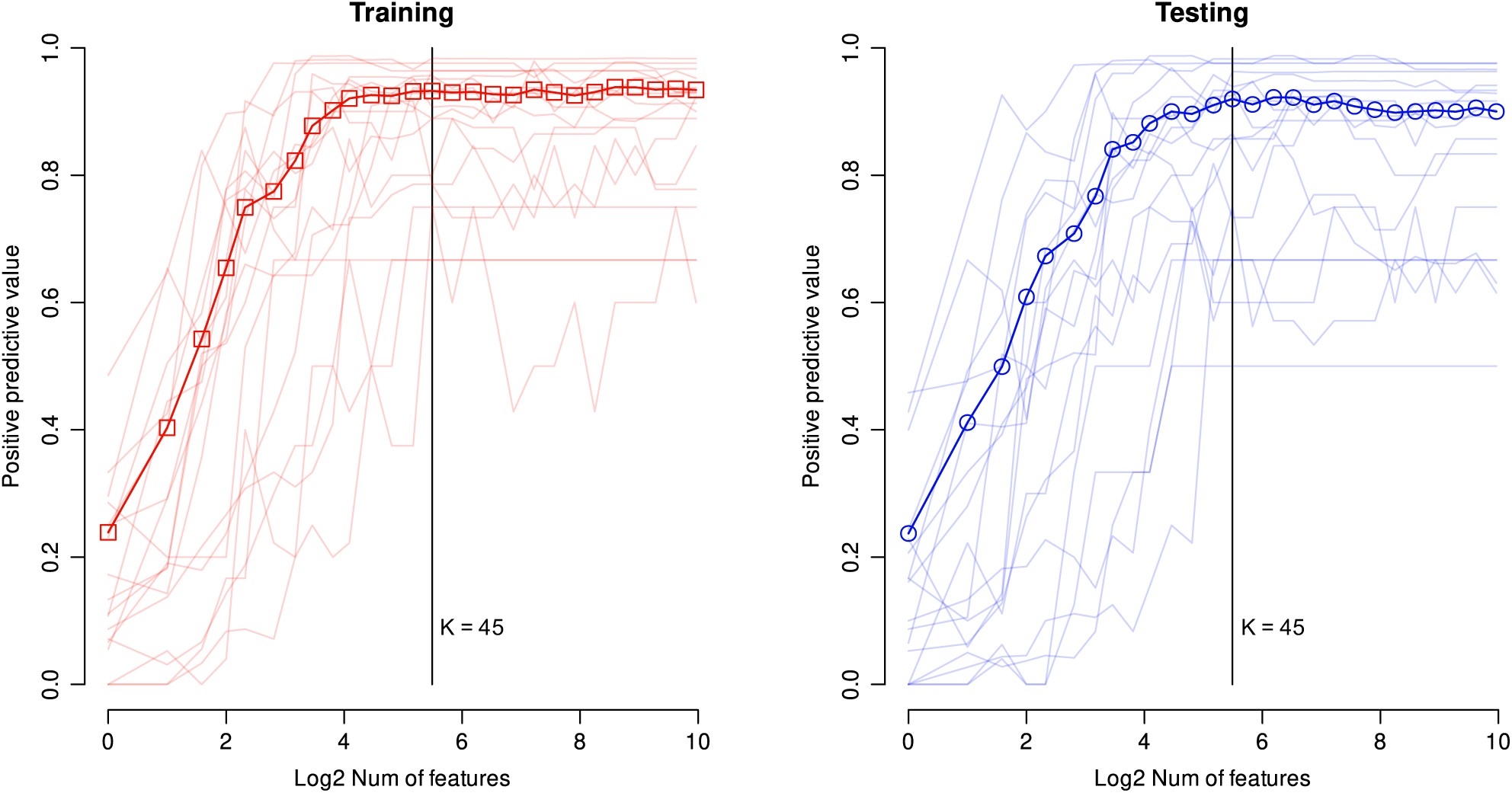
Optimization of a hyperparameter (number of input genes) for the centroid classifier. The model was trained with half the batch corrected North American dataset and tested with the other half. The curves depict overall positive predictive values that were calculated by applying the centroid models with different numbers of inputs to unadjusted North American data. We chose K = 45 as the final hyperparameter value.

**Supplement Figure S10.**
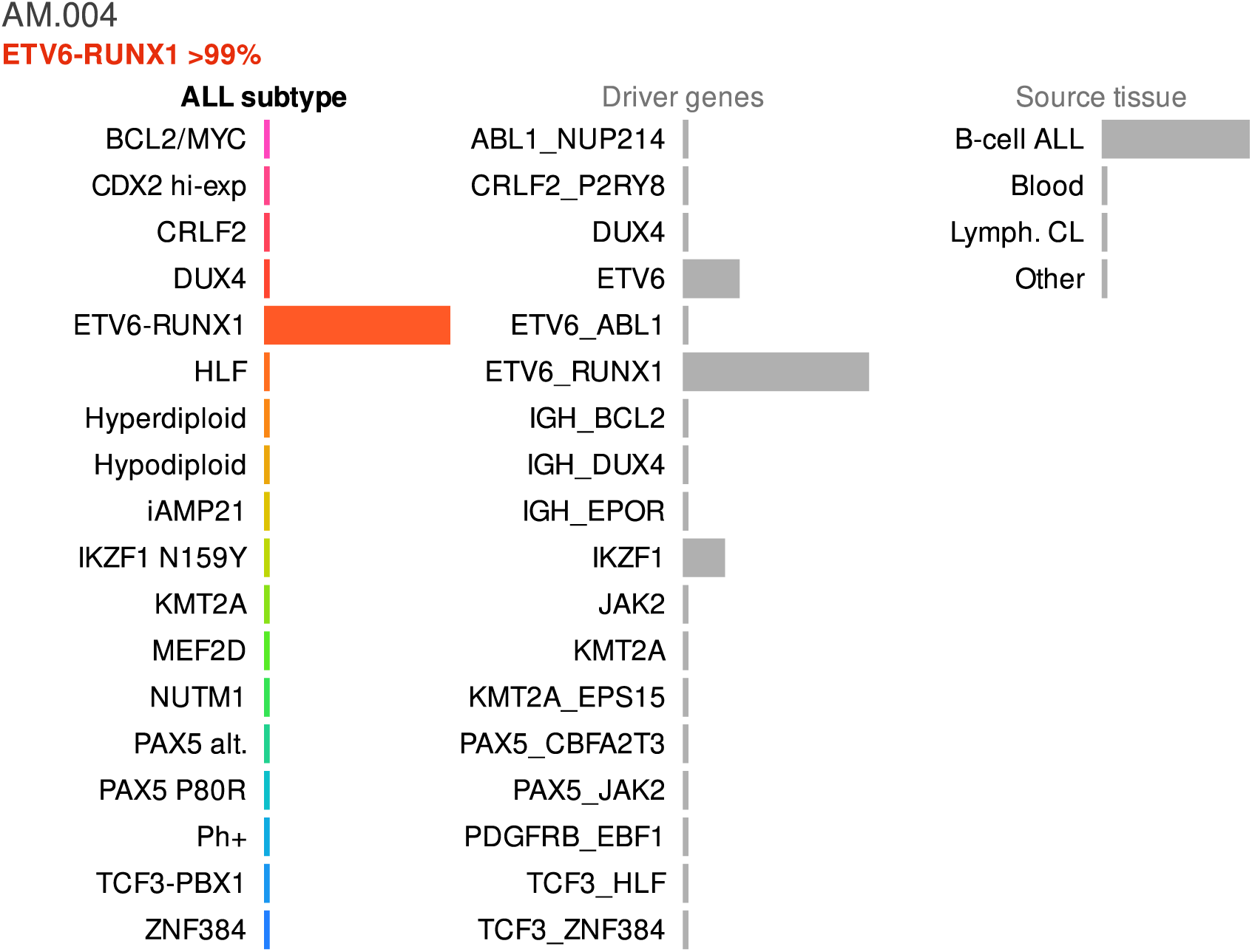
ETV6-RUNX1 case study from a North American cohort. This is an example of a patient with a distinct gene expression profile associated with a directly observable driver fusion.

**Supplement Figure S11.**
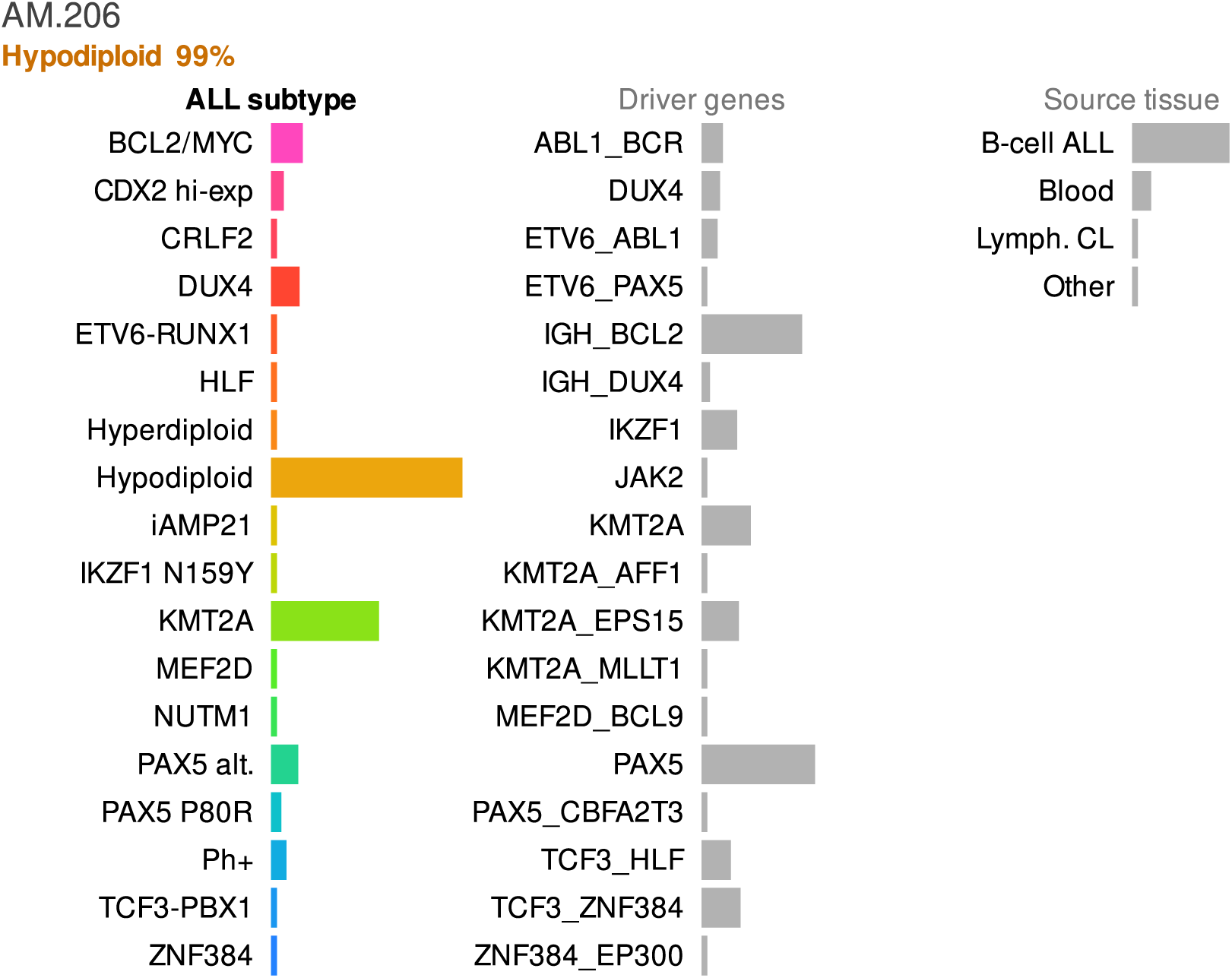
Hypodiploid case study from the North American dataset. Patients with chromosomal alterations are easy to detect via karyotyping, however, the extensive genetic alterations may affect multiple genes and pathways which may make it difficult to ascertain specific targets for molecular therapies. In this example, the RNA-seq profile suggests that the transcriptional consequences are compatible with a broad group of individuals that have BCL2, KMT2A and/or PAX5 lesions. The additional information from RNA-seq provides clues on the combination of genomic drivers that may be specific to this patient.

**Supplement Figure S12.**
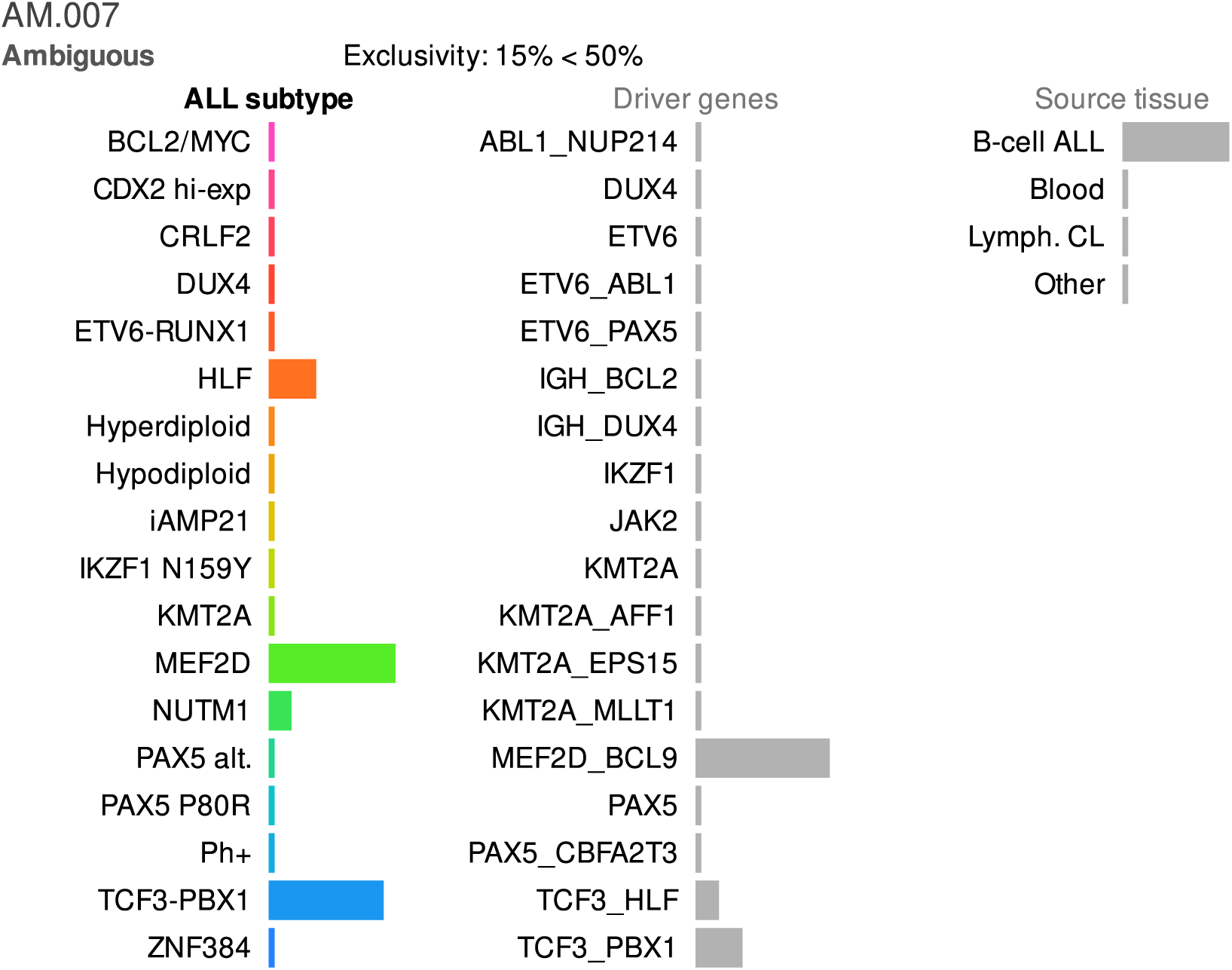
Ambiguous case study from the North American dataset. It is possible for a patient to exhibit multiple genetic lesions that drive ALL. Furthermore, the transcriptional consequences from different lesions may converge to similar profiles, which means that some patients will exhibit mixed gene expression characteristics. In this example, the transcriptional profile is in between the MEF2D and TCF3-PBX1 subtypes. The secondary analysis of driver genes (middle panel) suggests the combined lesion of MEF2D and BCL9 may be an important factor for this individual.

**Supplement Figure S13.**
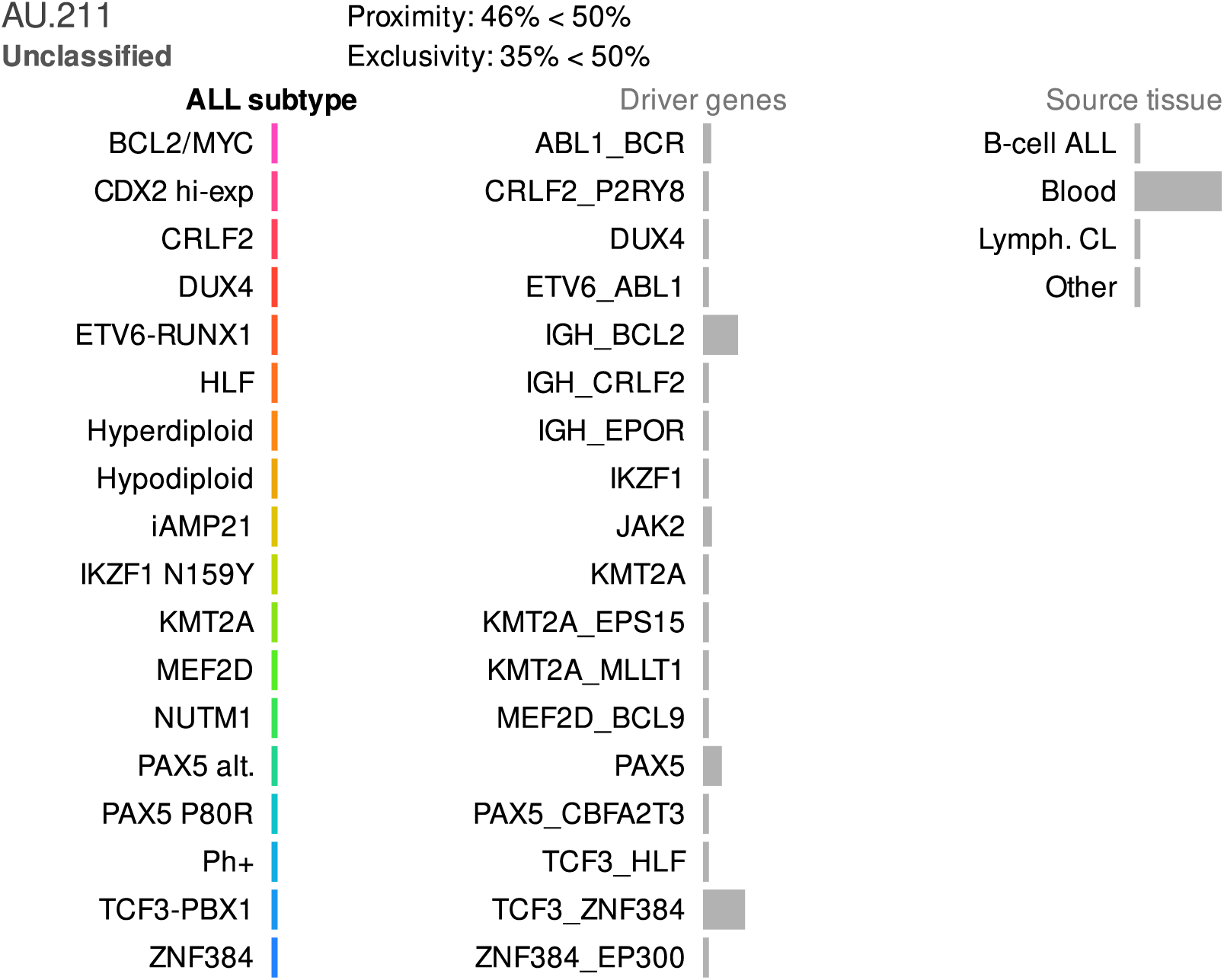
Unclassified case study from the Australian dataset. An unusual gene expression profile can represent a previously unknown subtype, however, it is more likely that the patient either had low leukemia burden or that incidental factors such as instrument failures or sample handling accidents had affected the sample quality. In Allspice, atypical samples are characterised by the lack of signals across the subtypes and genetic drivers. Here, there is also indication that leukemia burden may be low (see the tissue panel on the right).

**Supplement Figure S14.**
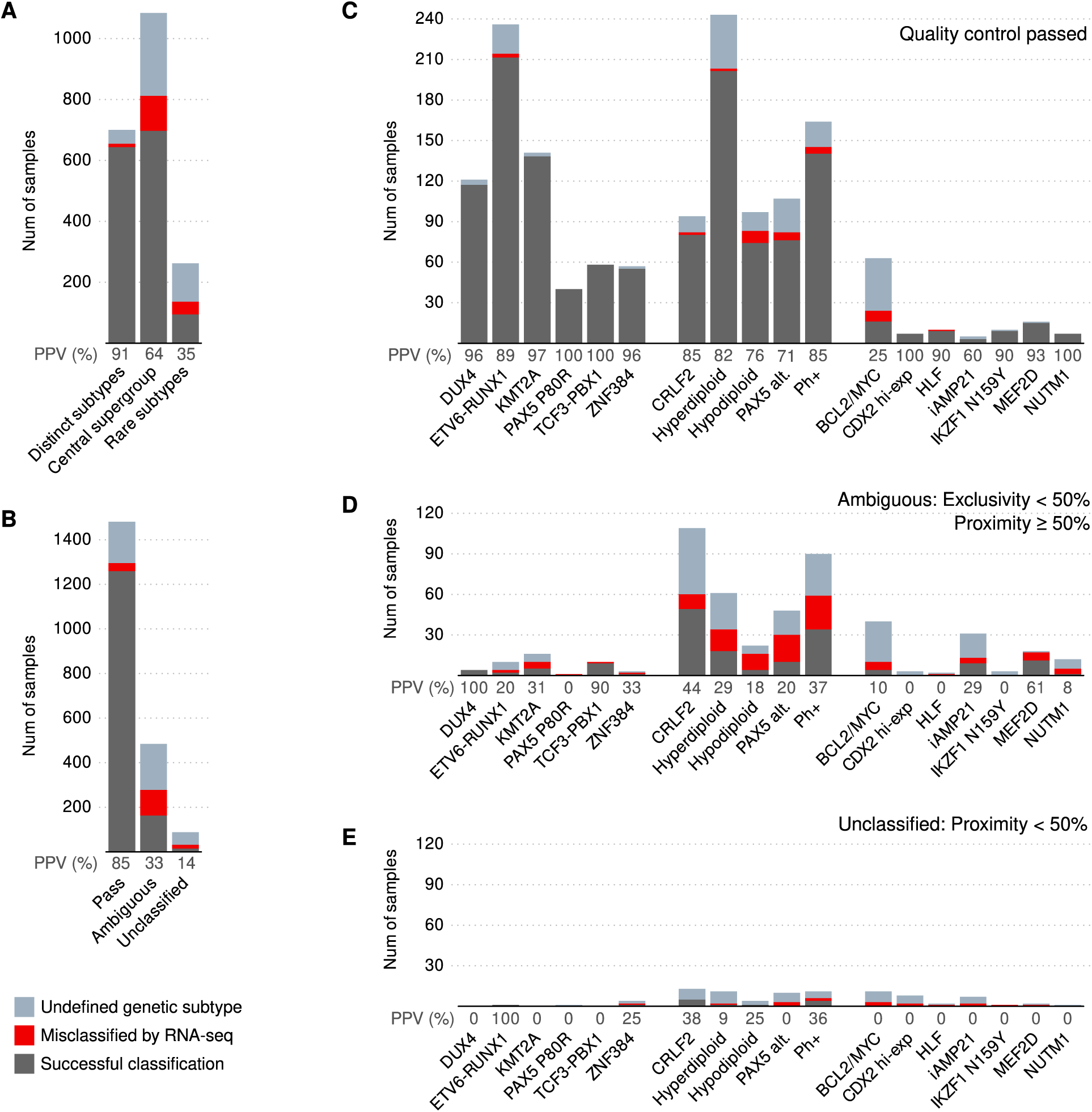
Detailed breakdown of the Allspice classifier performance with respect to subtypes and sample quality. **A**) Samples classified as one of the distinct subtypes exhibited high accuracy and a low proportion of undefined genetic subtypes. The central subgroup was less clear with lower accuracy overall. **B**) Samples that passed quality control (proximity ≥ 50%) were accurately classified. Ambiguous and unclassified samples included a high proportion of undefined subtypes. **C-E**) Samples classified by Allspice into specific subtypes, stratified into quality groups.

**Supplement Figure S15.**
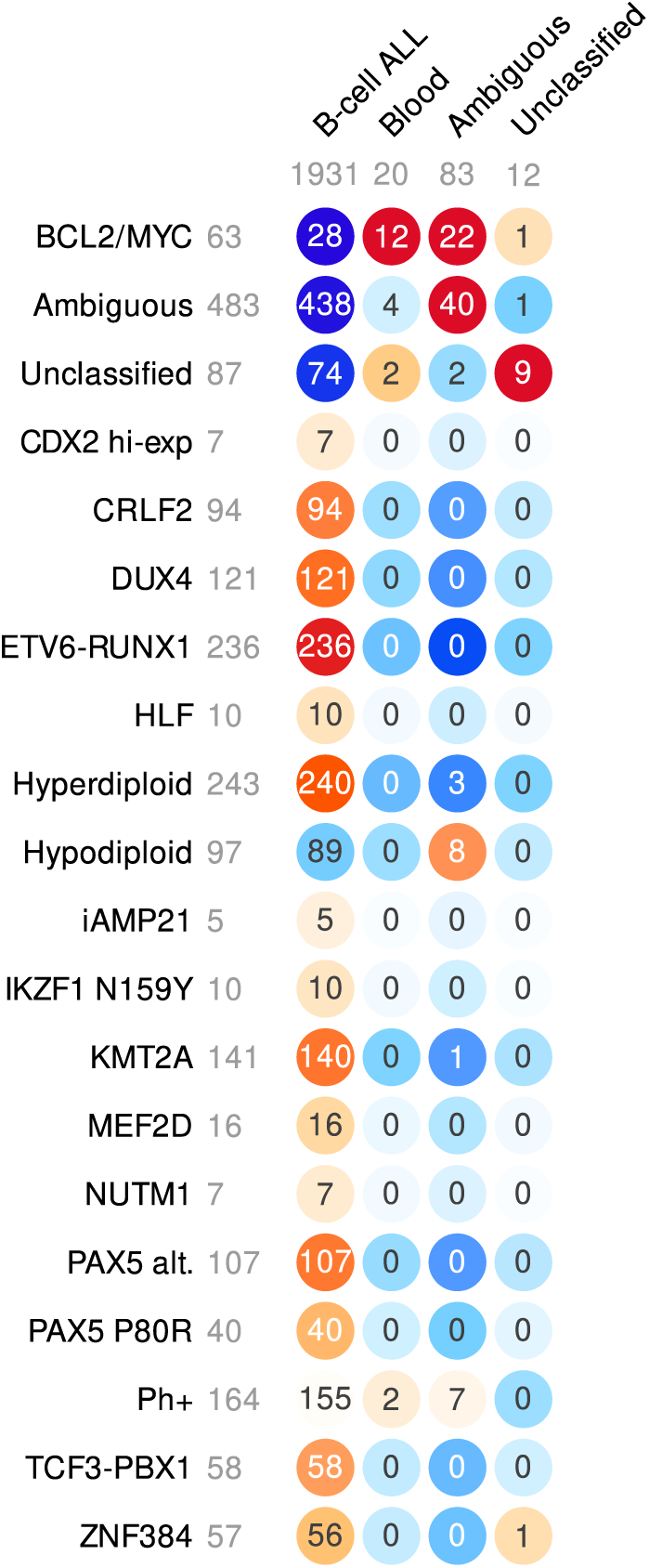
Application of a secondary tissue classifier contained within the Allspice package. The numbers of B-cell ALL samples are shown (total 2,046). The color intensity indicates deviation from the expected category distribution given the category sizes. Substantial classification of the ALL patient samples as whole blood or ambiguous source were observed for the BCL2/MYC subtype (35 out of 63).

**Supplement Figure S16.**
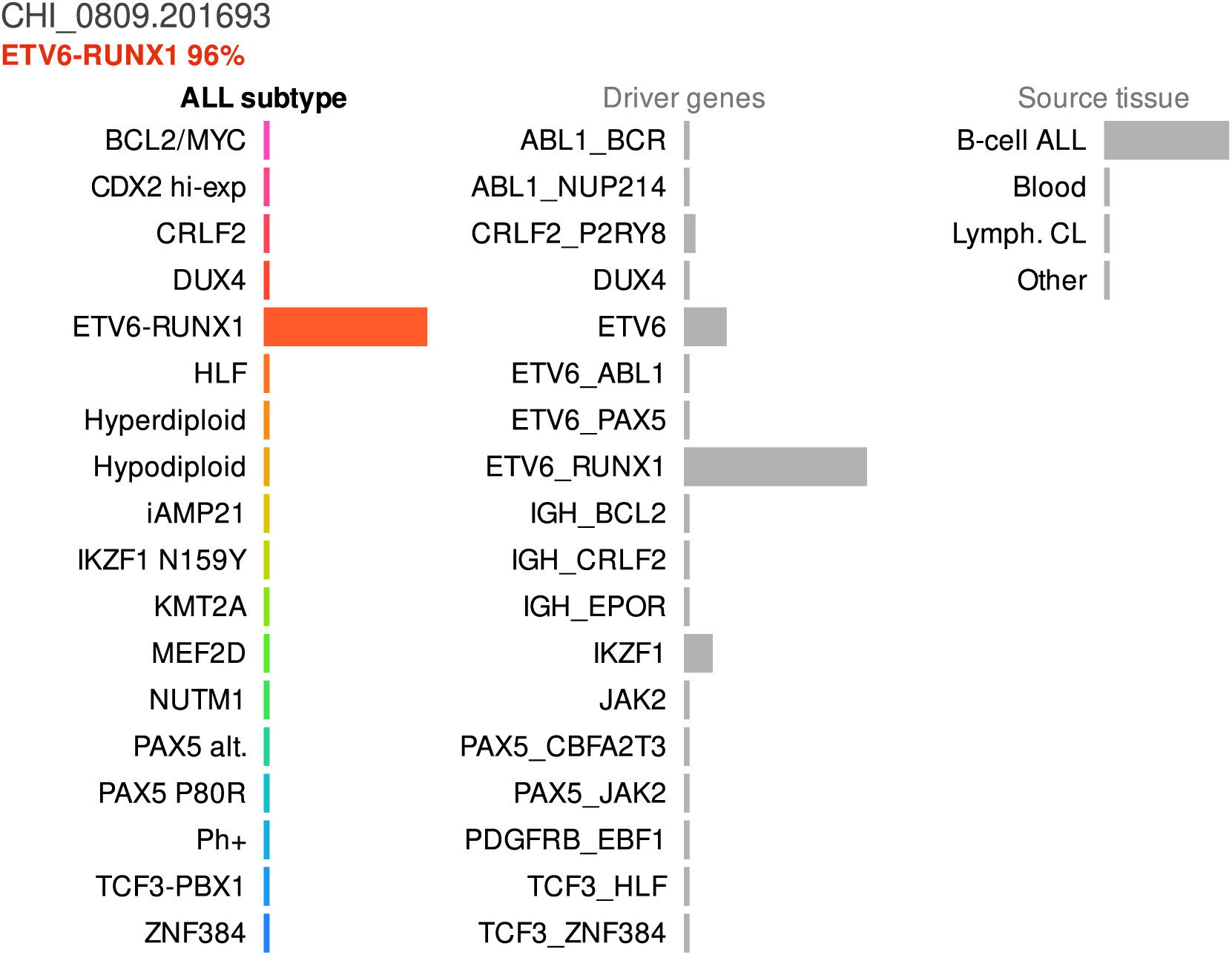
A case study of a patient who was tested at our study center in South Australia. The sample arrived after Allspice was finished and it was not used anywhere else in this manuscript. No ETV6-RUNX1 fusion was identified by molecular genetics (Karyotype, FISH, SNParray) or within the RNA-seq data. RNA-seq identified the following fusions (appear to be consistent with complex 3-way translocation idicated by FISH): ETV6 (chr 12, exon 7) - HDAC9 (chr 7, exon 13), HDAC9-ETV6 (3’ UTR exon 13 - intron 1), UBE4B (chr 1, exon 2) - ETV6 (chr 12, exon 8), ETV6 (chr 12, exon 8) - IKZF1 (chr 7, exon 3). However, the transcriptional profile was consistent with ETV6-RUNX1 and the patient was subsequently classified as ETV6-RUNX1-like.

**Supplement Figure S17.**
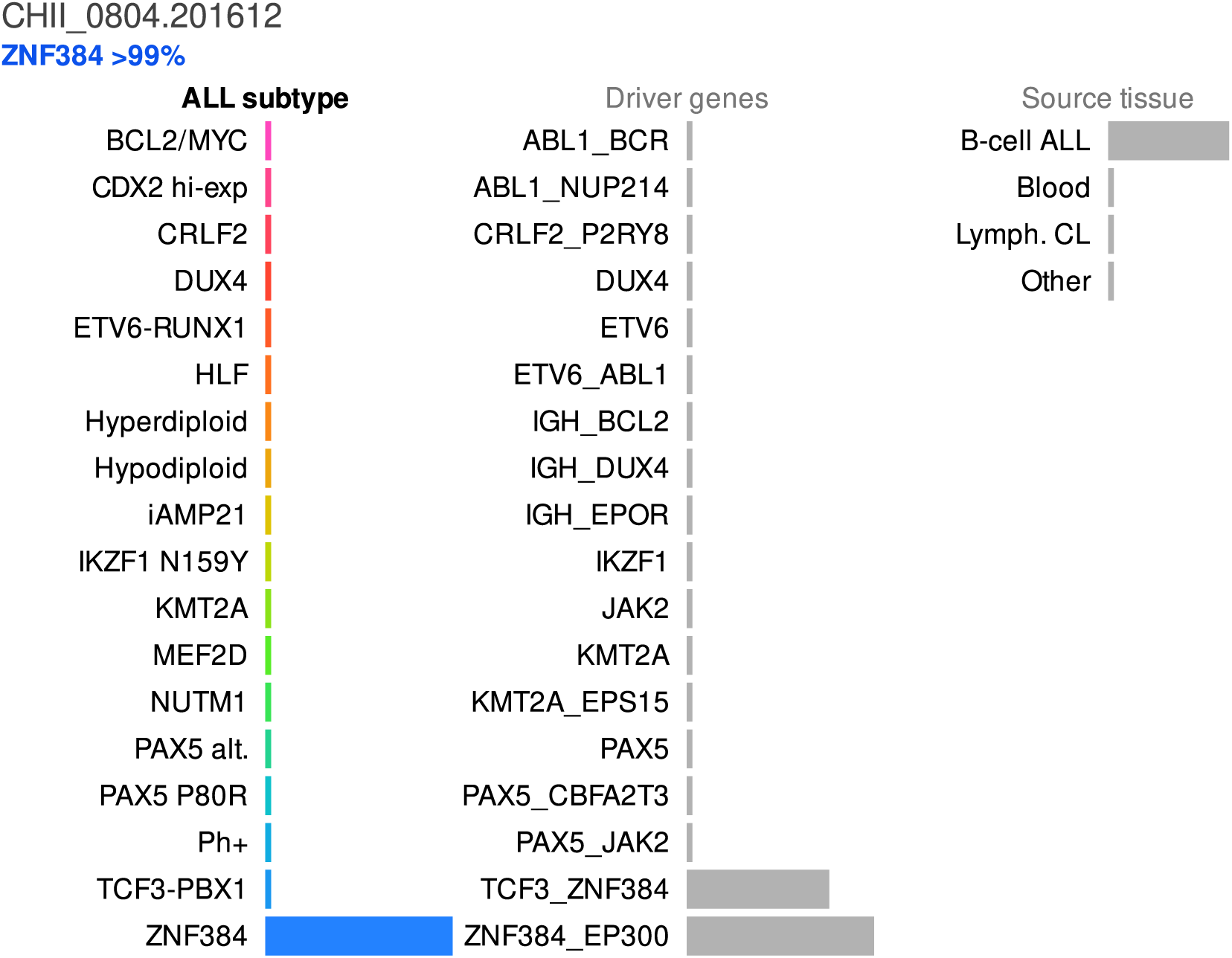
A case study of a patient who was tested at our study center in South Australia. The sample arrived after Allspice was finished and it was not used anywhere else in this manuscript. No known ZNF384 fusions identified by molecular genetics or RNA-seq. FISH performed with ZNF384 Break Apart Probe, no rearrangement found. Detailed search of sequence data identified an AHSA2-ZNF382 fusion called by 9 reads by fusion catcher only. In UCSC Genomic coordinates maps to intron 78 of USP34 and the intragenic region between ZNF383 and ZNF461. The patient was classified as ZNF384-like based on the transcriptional phenotype.

